# *PKHD1L1*, A Gene Involved in the Stereocilia Coat, Causes Autosomal Recessive Nonsyndromic Hearing Loss

**DOI:** 10.1101/2023.10.08.23296081

**Authors:** Shelby E. Redfield, Pedro De-la-Torre, Mina Zamani, Hanjun Wang, Hina Khan, Tyler Morris, Gholamreza Shariati, Majid Karimi, Margaret A. Kenna, Go Hun Seo, Hongen Xu, Wei Lu, Sadaf Naz, Hamid Galehdari, Artur A. Indzhykulian, A. Eliot Shearer, Barbara Vona

**Author notes:** Contributed equally to this work (Shelby E. Redfield, Pedro De-la-Torre). Contributed equally to this work (Mina Zamani, Hanjun Wang). Contributed equally to this work (Artur A. Indzhykulian, A. Eliot Shearer, Barbara Vona).

## Abstract

Identification of genes associated with nonsyndromic hearing loss is a crucial endeavor given the substantial number of individuals who remain without a diagnosis after even the most advanced genetic testing. *PKHD1L1* was established as necessary for the formation of the cochlear hair-cell stereociliary coat and causes hearing loss in mice and zebrafish when mutated. We sought to determine if biallelic variants in *PKHD1L1* also cause hearing loss in humans.

Exome sequencing was performed on DNA of four families segregating autosomal recessive nonsyndromic sensorineural hearing loss. Compound heterozygous p.[(Gly129Ser)];p.[(Gly1314Val)] and p.[(Gly605Arg)];p[(Leu2818TyrfsTer5)], homozygous missense p.(His2479Gln) and nonsense p.(Arg3381Ter) variants were identified in *PKHD1L1* that were predicted to be damaging using *in silico* pathogenicity prediction methods. *In vitro* functional analysis of two missense variants was performed using purified recombinant PKHD1L1 protein fragments. We then evaluated protein thermodynamic stability with and without the missense variants found in one of the families and performed a minigene splicing assay for another variant. *In silico* molecular modelling using AlphaFold2 and protein sequence alignment analysis were carried out to further explore potential variant effects on structure. *In vitro* functional assessment indicated that both engineered PKHD1L1 p.(Gly129Ser) and p.(Gly1314Val) mutant constructs significantly reduced the folding and structural stabilities of the expressed protein fragments, providing further evidence to support pathogenicity of these variants. Minigene assay of the c.1813G>A p.(Gly605Arg) variant, located at the boundary of exon 17, revealed exon skipping leading to an in-frame deletion of 48 amino acids. *In silico* molecular modelling exposed key structural features that might suggest PKHD1L1 protein destabilization.

Multiple lines of evidence collectively associate *PKHD1L1* with nonsyndromic mild-moderate to severe sensorineural hearing loss. *PKHD1L1* testing in individuals with mild-moderate hearing loss may identify further affected families.

## Introduction

Hearing loss-associated genes are implicated in the function of all parts of the delicate molecular machinery that permits human hearing. The inner hair cells (IHCs) and outer hair cells (OHCs) of the organ of Corti contain an apical bundle of ∼100 actin-filled protrusions called stereocilia. Upon sound stimulation, stereocilia bundles are deflected by pressure-induced waves within the fluid-filled organ of Corti. Housing the mechanotransduction apparatus at the tips of stereocilia, these bundles mediate the transformation of the mechanical stimulus into an electrical signal the brain interprets as sound. While IHCs convert sound waves into nerve signals, OHCs allow for non-linear amplification of the sound stimuli by changing their length in response to bundle deflection, a process known as electromotility (Brownell 1990). Although IHCs and OHCs have two separate and distinct functions, both sensory cell types require a properly organized, functional stereocilia bundle. Stereocilia have a transiently expressed surface coat that was first observed in the 1980s as an electron dense material (Santi and Anderson 1987; Slepecky and Chamberlain 1985), but little is understood about the function or molecular architecture of this surface specialization. To date, there are over 30 genes reported to be critical for stereocilia bundle morphology that are associated with sensorineural hearing loss (SNHL) in humans (Michalski and Petit 2015; Petit and Richardson 2009).

One such stereocilia protein, polycystic kidney and hepatic disease 1-like 1 (PKHD1L1), also called fibrocystin-L, is critical for hearing in mice (Wu et al. 2019). The *PKHD1L1* gene in humans encodes a 4,243 amino acid protein, which is predicted to be composed by a large extracellular domain, a 20 amino acid transmembrane domain, and a very short cytoplasmic domain of 8 residues. In mice, PKHD1L1 is highly enriched in both IHCs and OHCs, particularly at the tips of OHC stereocilia bundles (Wu et al. 2019). It is hypothesized that this protein makes up the majority of the transient stereocilia coat observed on the surface of hair cell stereocilia membrane. Mice lacking *Pkdh1l1* displayed elevated auditory brainstem responses (ABR) and distortion product otoacoustic emissions (DPOAE) thresholds in response to pure tone stimuli in a progressive fashion (Wu et al. 2019), and lacked the stereocilia coat. More recent data from zebrafish (*Danio rerio, Dr*) with a double knockout of *pkhd1l1a* and *pkhd1l1b* (orthologs of human (*Hs*) *PKHD1L1*) show significant deficits in auditory startle responses at the larval stage, consistent with an early-onset auditory phenotype in zebrafish (Makrogkikas et al. 2023). Based on these findings in animal models, we sought to determine whether variants in *PKHD1L1* cause hearing loss in humans.

In this study, we propose defects of *PKHD1L1* as causal for autosomal recessive nonsyndromic hearing loss in humans. We describe four unrelated families with biallelic variants in *PKHD1L1* identified via exome sequencing. All four probands presented bilateral congenital SNHL which is nonsyndromic and mild-moderate to severe. In addition, *in vitro* functional evaluation of two missense variants in protein fragments show decreased stabilities, suggesting that they may negatively impact their structures and molecular assembly *in vitro*, while a minigene assay of the c.1813G>A variant reveals aberrant splicing.

## Methods

### Recruitment and clinical assessment

This study was approved by the institutional review boards of Boston Children’s Hospital (IRB P-00031494), University Medical Center Göttingen (No. 3/2/16), and the School of Biological Sciences, University of Punjab, Lahore, Pakistan (IRB No. 00005281). Written informed consent was obtained from participating members of the four families or parents for their minor children.

The proband in family 1 derived from non-consanguineous parents and was ascertained as part of a cohort of 389 pediatric patients with SNHL in the United States. This cohort mostly includes individuals who were born to non-consanguineous parents (three probands with consanguinity reported). Any individual with SNHL was eligible for inclusion in the cohort regardless of SNHL laterality or severity, family history of SNHL, or presence of syndromic features. Two hundred forty-five probands had no prior genetic testing at the time of ascertainment, while 144 probands had some previous nondiagnostic genetic testing (variable methodologies). The proband in family 2 was derived from consanguineous parents who were first-degree cousins and was ascertained as part of a large ethnically diverse Middle Eastern population rare disease study consisting of approximately 800 probands with the sole inclusion criteria being hereditary hearing impairment. The proband in family 3 was born to consanguineous parents and identified from a special education school. The parents did not participate in the study. This proband is part of a cohort of 62 individuals with moderate to severe hearing loss who were born to consanguineous parents with no previous history of deafness in their families. The proband in family 4 was derived from non-consanguineous parents and sequenced as part of an Asian hearing loss cohort. This set is comprised of a total of 1450 hearing loss probands and it includes syndromic and non-syndromic hearing loss. Most of the cohort was first tested by a lab-developed multiplex PCR kit covering the total coding sequencing of *GJB2*, *SLC26A4*, and *MT-RNR1* (described in a previous study (Zeng et al. 2022b)), as these are the most common causative genes in this population. If negative, exome sequencing was performed (the so-called step-wise approach as described previously (Zeng et al. 2022b)). A fraction of patients chose exome sequencing as the first-tier test, while others with negative multiplex PCR were not tested by exome sequencing if DNA from both parents was not available. The proband in family 4 was identified from a subset of 449 probands for whom exome sequencing data were available.

Demographic, otolaryngologic, audiological, and relevant medical data were ascertained from the medical records of probands. Affected individuals underwent a complete otologic evaluation. Routine pure-tone audiometry was performed according to current standards in all probands and measured hearing thresholds at 0.25, 0.5, 1, 2, 4, 6, and 8 kHz. The probands in families 1 and 2 underwent tympanometry and speech audiometry testing, while the proband in family 4 underwent tympanometry. Probands 2 and 4 additionally underwent otoacoustic emissions testing. Pure-tone audiometry for proband 3 was performed in ambient noise conditions due to lack of soundproof testing environment.

### Exome sequencing

Genomic DNA (gDNA) from individuals in families 1 (I:1, I:2 and II:1), 2 ( I:1, I:2, II:1 and II:2), 3 (II:1), and 4 (I:1, I:2 and II:1) was isolated from either buccal mucosa or whole blood using standard procedures.

*Family 1:* Exome sequencing was performed in a Clinical Laboratory Improvements Amendments (CLIA)-certified environment (GeneDx, Gaithersburg, MD, USA). Analysis was performed using the DRAGEN pipeline (Illumina, San Diego, CA, USA). Copy number variants (CNVs) were called using the DRAGEN CNV pipeline and a normalized segmented depth of coverage model, as previously described (Rockowitz et al. 2020).

*Family 2*: Exome sequencing was applied to the DNA sample of the proband by Macrogen. Briefly, the sample was subjected to exome enrichment with the SureSelect Target Enrichment v6 kit (Agilent Technologies, Santa Clara, CA, USA), followed by sequencing with Illumina NovaSeq 6000 (Illumina, San Diego, CA, USA) using standard protocols. Then, short reads were aligned to the human genome reference version B38 using BWA and duplicate reads were marked using Picard. GATK and ANNOVAR were used for variant detection and annotation, respectively. Variant filtering and assessment was performed as previously described with slight modifications as described in the variant validation and assessment section (Vona et al. 2021).

CNVs were called using a read-depth based in-house tool, including exomeCopy (Love et al. 2011) and exomeDepth R packages (Plagnol et al. 2012). CNVs were predicted using a model of the normalized read depth.

*Family 3:* Exome sequencing for the proband was carried out at 3billion, Inc., Seoul, South Korea (https://3billion.io/index). Briefly, coding exon regions of human genes (∼22,000) were captured by xGen Exome Research Panel v2 (Integrated DNA Technologies, Coralville, IA, USA). The captured regions of the genome were sequenced with NovaSeq 6000 (Illumina, San Diego, CA, USA). The raw genome sequencing data analysis, including alignment to the GRCh37/hg19 human reference genome and variant calling and annotation, were conducted with open-source bioinformatics tools including Franklin (https://franklin.genoox.com/clinical-db/home) and 3billion in-house software.

*Family 4:* Exome sequencing, bioinformatics analysis, and variant filtering for the proband were performed as previously described (Zeng et al. 2022b). Copy number analysis was performed using DECoN (Fowler et al. 2016) with default settings and the BAM files from the same enrichment panel and sequencing run as controls. *STRC* copy number testing was performed using a PCR with exon 22 specific primers as (Vona et al. 2015) previously described, with MLPA analysis.

### Variant assessment and validation

All exome datasets were assessed without a retrospective search to find *PKHD1L1* biallelic variants. Exome data for the sequenced individuals in families 1, 2, and 3 was filtered to remove all variants with an allele frequency of 0.01 or more in public databases. Coding and splice site variants were retained. Deleteriousness of the missense variants was assigned according to prediction from multiple software and supported by evolutionary conservation of the affected amino acid (Table 1). Due to pedigree structures, homozygous variants were considered first while heterozygous variants were observed later. Additionally, variant analysis of the proband in family 2 employed an in-house database (including more than 2,500 exome datasets). Variants with low allele frequency and deleterious prediction were classified using the hearing loss-adapted ACMG criteria (Oza et al. 2018) and prioritized for further allele segregation studies in the family.

**Table 1.**
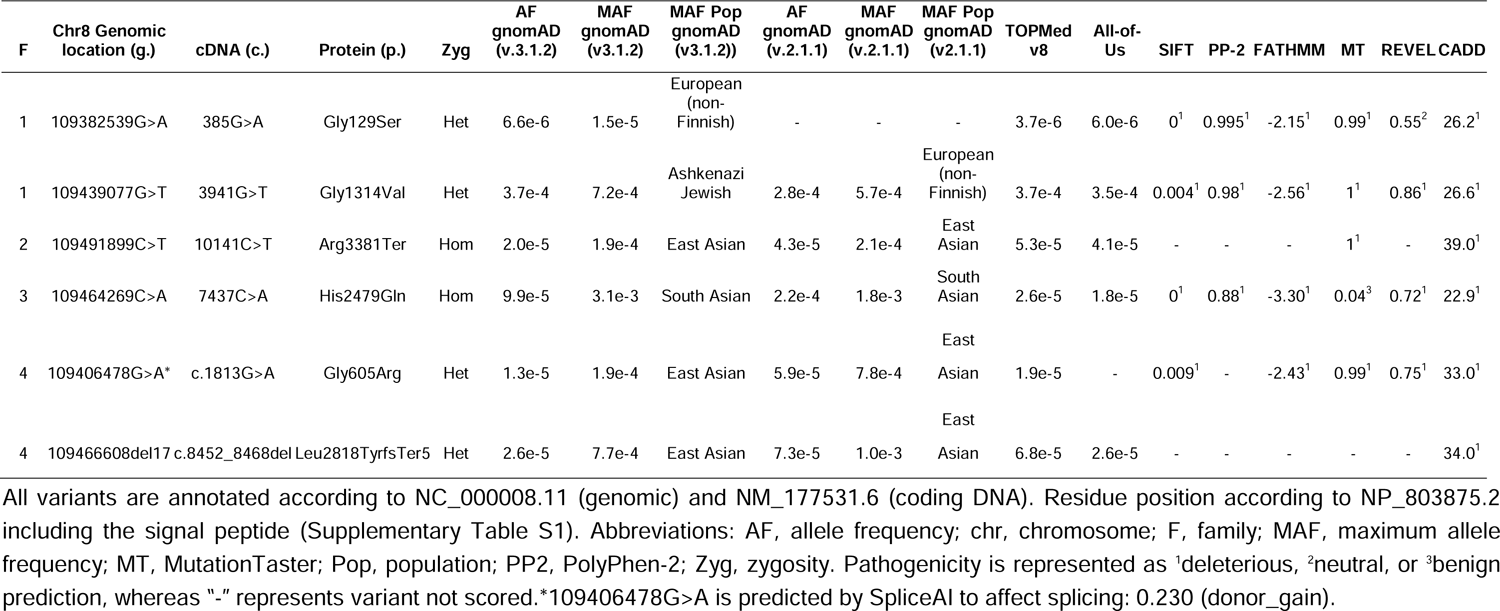
*PKHD1L1* variants identified.

Variants in the exome data of proband in family 4 were identified with SnpEff annotation. First, the following variant types were removed from the analysis: intergenic_region, upstream_gene_variant, downstream_gene_variant, 5_prime_UTR_variant, 3_prime_UTR_variant, intron_variant, and non_coding_transcript_exon. At the same time, variants with ada_score >0.5 or rf_score >0.5 and variants annotated as likely pathogenic, pathogenic, or VUS in ClinVar were retained. We then filtered out variants with minor allele frequency >0.001 in any population in which at least 2,000 alleles were observed in the gnomAD database (v2.1.1), except those on the ACMG benign stand-alone exception list. We prioritized variants that occurred in the curated hearing loss associated gene list from the ClinGen Hearing Loss Gene Curation Expert Panel (DiStefano et al. 2019).

Variants were prioritized based on population and *in silico* pathogenicity software predictions. Variant minor allele frequencies were derived from gnomAD (v2.1.1 and v3.1.2, Table 1) (Chen et al. 2022; Karczewski et al. 2020). Various pathogenicity prediction tools were used including SIFT (Ng and Henikoff 2001), PolyPhen-2 (Adzhubei et al. 2010), FATHMM (Shihab et al. 2014), MutationTaster (Schwarz et al. 2014), REVEL (Ioannidis et al. 2016) and CADD (Rentzsch et al. 2019). Variants were analyzed for splice prediction using SpliceAI (Jaganathan et al. 2019), and visualization of amino acid substitution tolerance was supported by the MetaDome web server (Wiel et al. 2019).

Variants were annotated using the *PHKD1L1* NM_177531.6 accession number (ENST00000378402.9). The GTEx Portal (Consortium 2013) was referenced for assessing the location of variants across the annotated *Hs PHKD1L1* gene sequence (Supplementary Fig. S1). *PKHD1L1* variant segregation in families 1, 2, and 4 was confirmed using Sanger sequencing, but not for the proband in family 3.

### Sequence analyses and structural modeling of PKHD1L1 protein

We compared PKHD1L1 protein sequences among 10 different PKHD1L1 orthologs (NM_177531.6 and NP_803875.2 for *Hs* PKHD1L1, see Supplementary Table S1 for more details about the selected species). The sequences were obtained from the National Center for Biotechnology Information (NCBI) protein database (see Supplementary Table S1 for NCBI accession numbers). First, each individual protein sequence was used to predict their signal peptides and domains using the Simple Modular Architecture Research Tool (SMART) (Letunic et al. 2021). Signal peptides were further predicted using the SignalP-5.0 (Almagro Armenteros et al. 2019) and the Prediction of Signal Peptides (PrediSi) online servers (Hiller et al. 2004). AlphaFold2 modelling was used to predict the potential signal peptide cleavage site and accurately inform the start and end of each predicted domain before carrying out the protein sequence alignment (Mirdita et al. 2022). Since the PKHD1L1 Ig-like-plexins transcription factors (IPT) domains do not have a clear conservation pattern at their IPT protein start and end sequence and connecting linker domains, AlphaFold2 modeling results were combined with protein sequence alignment to better predict the signal peptide, IPT domain start and end residue positions, and the location of missense mutations.

The ClustalW algorithm (Larkin et al. 2007) on Geneious (Kearse et al. 2012) was used for the sequence identity analysis using the default parameters. Alignment files from Geneious were imported and color-coded in JalView with 35% conservation threshold, as previously described (Kearse et al. 2012). AlphaFold2 simulations of PKHD1L1 fragments were carried out using the Colabfold v1.5.2-patch server using default parameters (Mirdita et al. 2022).

### Cloning, expression, and purification of engineered bacterially expressed PKHD1L1 protein fragments and mutant constructs

The cDNA of wild-type (WT) *Mus musculus* (*Mm*) *PKHD1L1* IPT1-3 and IPT5-6 were subcloned into the *Nde*I and *Xho*I sites of the pET21a+ vector. Next, the cDNA fragments were amplified from longer synthetized sequences optimized for *E. coli* expression. Site-directed mutagenesis was applied to engineer the *Mm* PKHD1L1 IPT1-3 p.(Gly129Ser) and PKHD1L1 IPT5-6 p.(Gly1314Val) constructs using the QuickChange lightning kit (Agilent Technologies). All constructs were used for protein expression in Rosetta (DE3) competent cells (Novagen) and cultured in TB as reported previously (De-la-Torre et al. 2018). Expressed recombinant proteins were purified under denaturing conditions (6 M guanidine) using nickel beads. Next, their purity was analyzed by sodium dodecyl sulfate-polyacrylamide gel electrophoresis (SDS-PAGE) and refolded at 4 °C using previously reported conditions (De-la-Torre et al. 2018), briefly outlined below. WT *Mm* PKHD1L1 IPT1-3 and IPT1-3 p.(Gly129Ser) were refolded by fast or drop-wise dilution as previously reported for other protein families (De-la-Torre et al. 2018): 20 mL of pure denatured protein (0.5-1 mg/mL) was dropped into 480 mL of refolding buffer containing 20 mM TrisHCl [pH 8.0], 150 mM KCl, 50 mM NaCl, 2 mM CaCl_2_, 400 mM L-arginine, and 2 mM *D*(+) glucose. WT *Mm* PKHD1L1 IPT5-6 and IPT5-6 p.(Gly1314Val) were refolded by dialysis of 40 mL of eluted denatured protein at 0.5 mg/mL into 1000 mL of refolding buffer (20 mM TrisHCl [pH 7.5], 150 mM KCl, 50 mM NaCl, 5 mM CaCl_2_, 400 mM *L*-arginine, 1 mM of glutathione oxidized). Proteins were concentrated using 10,000 Da Amicon Ultra-15 centrifugal filters and purified using size exclusion chromatography (SEC) with an Akta Purifier System with the S200 16/600 pg and S200 13/300 increase GL columns (GE Healthcare) in a buffer containing 20 mM Tris-HCl pH 7.5, 150 mM KCl, 50 mM NaCl, and 5 mM CaCl_2_ to preserve the most abundant endolymphatic cations. Following SEC, protein purity was further verified by SDS-PAGE.

### Nanoscale differential scanning fluorimetry (NanoDSF)

WT *Mm* PKHD1L1 IPT1-3 and IPT5-6 protein fragments and their respective missense IPT1-3 p.(Gly129Ser) and IPT5-6 p.(Gly1314Val) proteins were used for functional evaluation *in vitro*. Thermodynamic evaluation and folding stabilities of these constructs in solution were carried out using nano differential scanning fluorimetry (NanoDSF). Pure proteins were concentrated to 0.5-1 mg/mL for NanoDSF using a Prometheus NT.48 (Nanotemper) and scanned between 20-95°C with a pre-stabilization phase of 1 min and a temperature slope of 2 °C/min (37 min in total). Data were processed using a PR. ThermControl v2.1.2 software and plotted using GraphPad Prism. At least two biological replicates were used for each experiment. Each protein preparation was independently expressed and refolded at least two time (two biological replicates), and evaluated independently on NanoDSF. Each NanoDSF scan used at least four separate capillary tubes run in parallel for each biological replicate. Each result per biological replicate represents average values of these measurements. Protein folding analysis results were plotted as a relationship of the normalized F350/F330 (%) ratio intensities (T_onset_). The first derivative of F350/F330 (%) intensities were plotted to obtain the thermal unfolding transition midpoints (T_m_).

### Minigene assay of the c.1813G>A (Gly605Arg) variant

A minigene assay was performed as previously described (Zeng et al. 2022a). Briefly, the WT and mutated sequences (exon 17 and flanking intronic sequences) were cloned (MINI-PKHD1L1-Kpn1-F: 5’-GGTAGGTACCAGGCC-3’, 5’-TATGGAACACCAATTTA-3’ and MINI-PKHD1L1-BamH1-R: 5’-TAGTGGATCCAAT-3’ and 5’-AAGGCCTGTCCTCAAATGTCT-3’) following amplification from the DNA of the proband in family 4 between exons A and B in the pcMINI plasmid. Next, the WT and mutated plasmids were transfected into both HEK293 and HeLa cells. The splicing effects were analyzed via RT-PCR and sequencing with vector-specific primers (PcMINI-F: 5’-CTAGAGAACCCACTGCTTAC-3’ and PcMINI-R: 5’-TAGAAGGCACAGTCGAGG-3’).

## Results

### Clinical genetics and variant identification

**Family 1 (Fig. 1):** The proband is a 11-15-year-old female born to healthy nonconsanguineous parents. She did not pass a newborn hearing screen bilaterally. Pure tone audiometry has been performed approximately every 6 months and consistently demonstrated a slowly progressive mild to moderate SNHL bilaterally. Between the ages of 0-5 years and 11-15 years, there was an increase in pure tone average (PTA) of 5 dB for the right ear and 8 dB for the left ear. PTA_0.5-4K_ was 41.25 and 38.75 in the right and left ears, respectively at age 0-5. Most recent audiometric testing showed PTA of 45.00 and 48.75 in the right and left ears, respectively. Speech audiometry at most recent evaluation (age 11-15 years) demonstrates a 90%-word recognition at a comfortable listening level. Speech recognition threshold (SRT) is 45 dB bilaterally. There was a history of episodes of benign paroxysmal positional vertigo (BPPV) which resolved after Epley maneuver. Imaging studies of the inner ear were not performed. An electrocardiogram and ophthalmology exam were normal. Cytomegalovirus testing performed at 11-15-weeks-old was negative. There were no dysmorphic facial features, neurological or developmental abnormalities, or other pertinent history. Exome sequencing was performed with an average depth of coverage of variants of 92 reads with 95.4% of variants covered with more than 20 reads. Exome sequencing revealed candidate variants only in *PKHD1L1* following the variant filtering methodology described. The proband was compound heterozygous for missense variants, c.385G>A, p.(Gly129Ser) and c.3941G>T, p.(Gly1314Val) (Table 1) that segregated within the trio in a Mendelian recessive manner (residue numbering corresponds to the NCBI *Hs* PKHD1L1 sequence NP_803875.2 including the signal peptide, Supplementary Table S1).

**Fig. 1.**
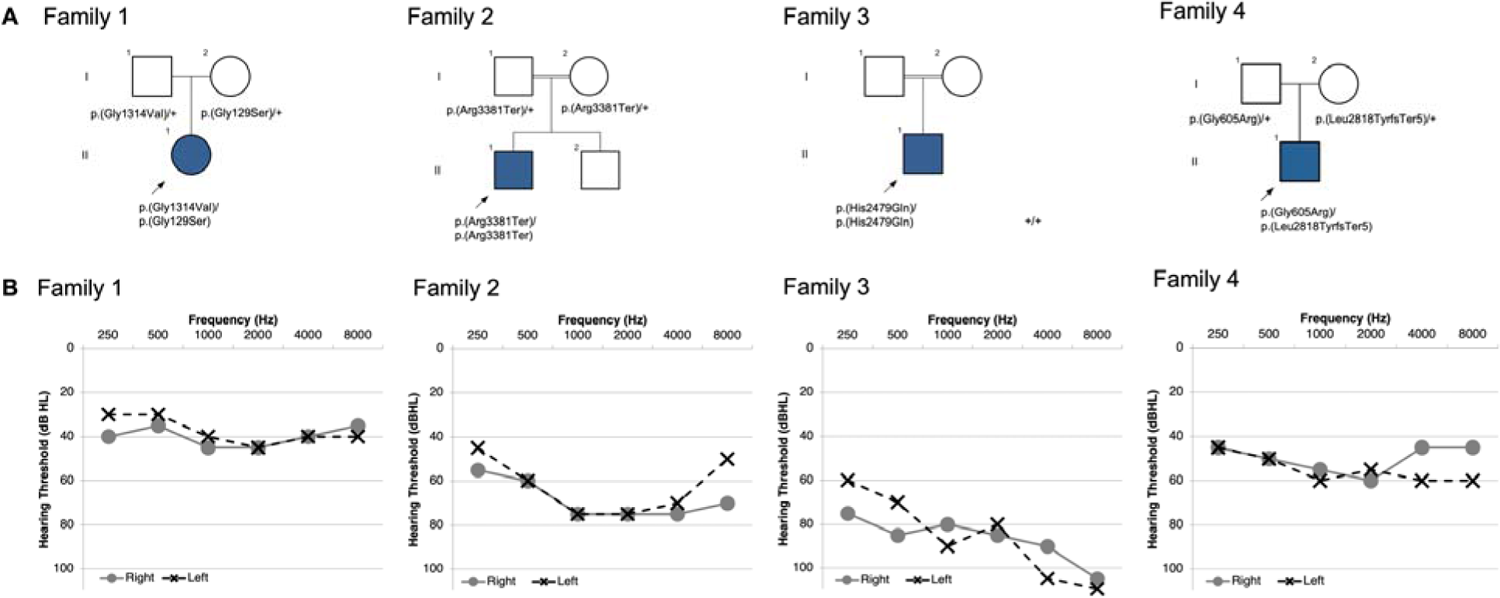
Pedigrees & Audiograms. Pedigree and audiometric information for four families with biallelic *PKHD1L1* variations (a) Pedigrees for families 1-4. Each proband with SNHL is indicated with shading and arrow. (b) Pure tone audiometry for probands 1-4; x represents results for the left ear and o represents the right. Audiometric evaluation performed at ages ranging between 8 and 13 years-old for probands.

The c.385G>A, p.(Gly129Ser) substitution resides in exon 4 of 78 in *PKHD1L1* and has a maximum allele frequency (MAF) of 0.001471% in gnomAD (v3.1.2). This variant is predicted to be deleterious to protein structure and function via *in silico* predictors (Table 1). This nonpolar glycine to polar serine substitution occurs at the tip of the PKHD1L1 IPT1 domain (N-terminal end) (Fig. 2b-d). This locus is predicted to be somewhat tolerant to missense substitution (Supplementary Fig. S2). On the other hand, the c.3941G>T, p.(Gly1314Val), located in exon 32 of 78 in *PKHD1L1,* has a MAF of 0.07204% (Table 1, gnomAD, v3.1.2). It is suggested to be deleterious to protein structure and function by *in silico* prediction tools (Table 1), as well as predicted to be intolerant to missense substitution (Supplementary Fig. S2). The p.(Gly1314Val) substitution is located at the PKHD1L1 IPT6 domain region (Fig. 2b-e).

**Fig. 2.**
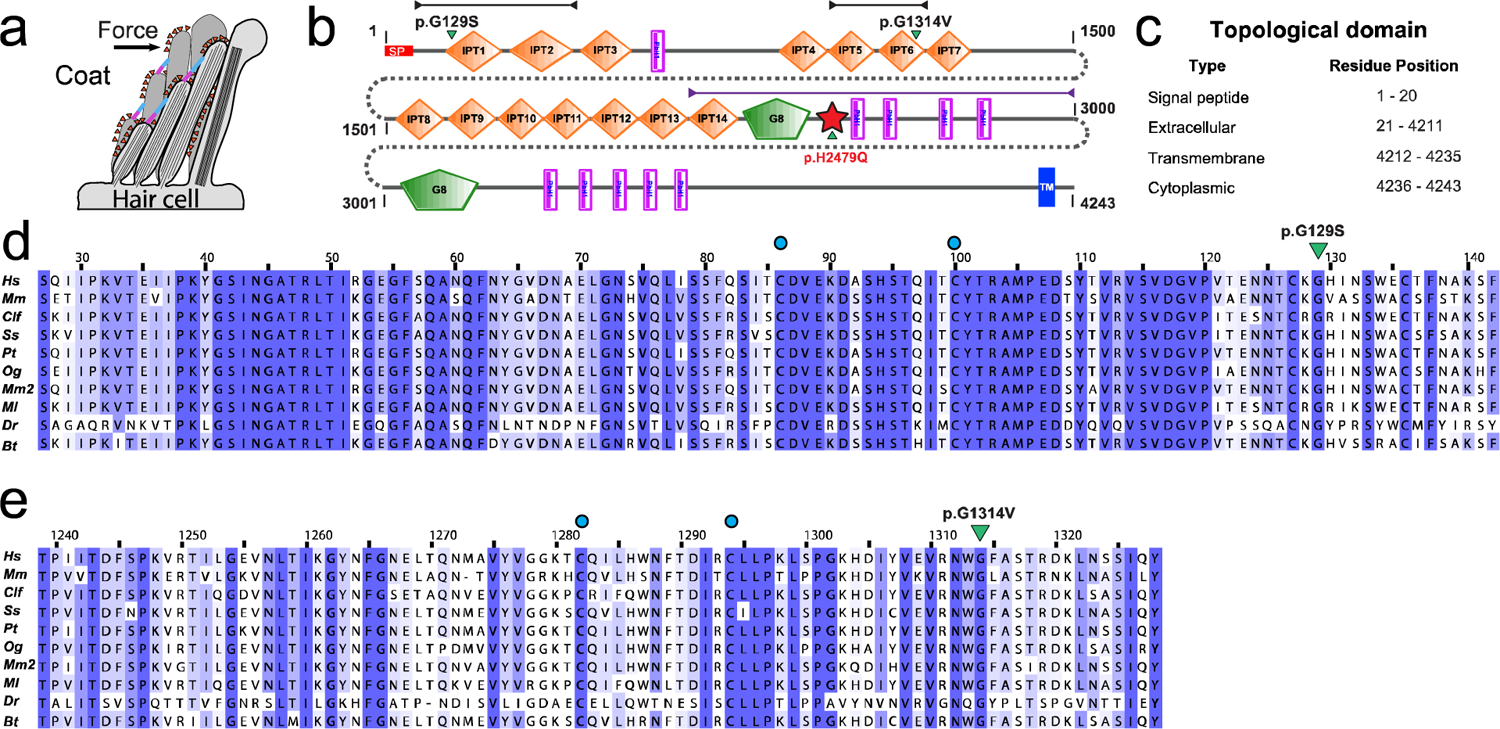
PKHD1L1 protein domain prediction and evolutionary analysis. (a) Schematic of a hair-cell stereocilia bundle under force stimulation highlighting the stereocilia surface coat. (b) Protein domain composition prediction from SMART using the *Hs* PKHD1L1 protein sequence as in NCBI accession code NP_803875.2, including the signal peptide (20 amino acids are predicted for *Hs* PKHD1L1 according to SMART. See Supplementary Table S1). Positions of each missense variant reported in this study is presented with a green arrowhead. The red star represents a newly predicted TMEM2-like domain. Black and purple arrow-headed lines represent the sequence fragments used for AlphaFold2 modelling of IPT1-2, IPT5-6, and TMEM2-like domain, respectively. (c) Topological description of *Hs* PKHD1L1 protein sequence as a reference. (d-e) Multiple protein sequence alignments comparing IPT1 and IPT6 domains among 10 different PKHD1L1 orthologs, respectively (see Supplementary Table S1 for details about the selected species and Supplementary Fig. S3 for full PKHD1L1 sequence alignment). IPT1 has a pairwise sequence identity conservation of 82.3%, while IPT6 has a pairwise sequence identity of 74.9% across 10 different orthologs. An independent % sequence identity analysis of only *Hs* and *Mm* species for IPT1 and IPT6 shows 82.9% and 77.8%, respectively (sequence alignment not shown). Missense variants are highlighted by green triangles. Blue circles represent cysteine residues forming disulfide bonds showed in figure 2 d-e. Each alignment was color-coded for sequence similarity (35% threshold) using Jalview. White-colored residues report the lowest similarity and dark blue report the highest (see Methods). PKHD1L1 orthologs were chosen based on sequence availability and taxonomical diversity (Choudhary et al. 2020; De-la-Torre et al. 2018; Jaiganesh et al. 2018).

**Family 2 (Fig. 1):** The proband is a 6-10-year-old male born to healthy consanguineous (first cousin) parents with subjectively normal hearing. Congenital SNHL was suspected that was clinically diagnosed in the first months of life and has progressed to a bilateral moderate to severe degree. Pure tone audiometry shows moderate to severe SNHL in all frequencies. Speech audiometry understanding is 100% at a comfortable listening level, and the otoacoustic emissions were present bilaterally. His SRT is 60 dB and his speech discrimination score (SDS) is 100% at the intensity level of 80 dB. The proband currently uses hearing aids bilaterally. There have been no vestibular abnormalities or delays in motor milestones. Exome sequencing was performed with an average depth of 66X, with 75% of variants covered with more than 20 reads. Exome sequencing revealed that the proband was homozygous for the c.10141C>T, p.(Arg3381Ter) nonsense variant (Table 1) that resided in a ∼28 Mb run of homozygosity. All other variants were excluded based on segregation (Supplementary Table S2). Sanger sequencing at this locus confirmed the homozygous variant and revealed that the parents were both heterozygous carriers of the c.10141C>T, p.(Arg3381Ter) substitution.

The c.10141C>T, p.(Arg3381Ter) variant is located in exon 62 of 78 and identified with a MAF of 0.02067% in population databases (Table 1, gnomAD, v2.1.1). This variant occurs in a region with parallel beta-helix (Pbh1) repeats (Fig. 2b and Supplementary Fig. S3 for residue conservation), introducing a premature stop codon that is predicted to result in the loss of approximately 20% of the transcript (∼ 882 amino acids), including the transmembrane domain, and could potentially cause nonsense mediated decay. However, if expressed in a truncated form, lack of the transmembrane domain is likely to impair proper localization of PKHD1L1 in the cell membrane or might induce unconventional secretion of PKHD1L1 protein fragments.

**Family 3 (Fig. 1):** The proband is a 11-15-year-old male with congenital SNHL born to healthy consanguineous parents, and her audiometric testing demonstrated a bilateral severe SNHL (PTA_0.5-4K_ 85 dB HL). No further follow up was possible for the affected individual. Exome data had an average overall depth of 211X and 99.1% of variants were covered by more than 20 reads. Exome analysis revealed two homozygous missense variants of interest: one in *PKHD1L1* c.7437C>A, p.(His2479Gln) and one in *MYO7A* (NM_000260.4:c.1123C>G, p.(Leu375Val)). Both *MYO7A* and *PKHD1L1* variants were of high quality and each was covered by more than 150 reads. The homozygous variant in *MYO7A*, with a coverage of 198 high quality reads, was deprioritized given uncertain and neutral *in silico* predictions with respect to impact on protein structure and function (Supplementary Table S2 and Supplementary Fig. S6). Moreover, the affected amino acid was not conserved in evolution, being isoleucine instead of leucine in some mammals, birds, and amphibians.

The c.7437C>A, p.(His2479Gln) substitution in *PKHD1L1* is located in exon 49 of 78 and is identified at a MAF of 0.3107% in population databases (Table 1, gnomAD, v3.1.2). This positively charged histidine to neutral glutamine substitution is located in a topological region with an unpredicted domain structure when using the SMART prediction tool (Fig. 2b).

**Family 4 (Fig. 1):** The proband is an 6-10-year-old boy with SNHL born to healthy non-consanguineous parents. He presented in 2023 with concern for hearing loss, and pure tone audiometry demonstrated a bilateral moderate SNHL. DPOAEs were absent in both ears, and the tympanograms were normal, suggesting dysfunction of the outer hair cells. The hearing loss in this proband is thought to be congenital because the father claimed that the hearing was affected from a younger age, but newborn hearing screening was not performed at the time of birth. The father had a pure tone audiometric evaluation at the age of 30-35 years, which showed thresholds within the normal range.

The proband underwent exome sequencing at Precision Medicine Center of Zhengzhou University. Exome sequencing was performed with an average depth of 123.7X for all variants, with 99.2% covered by more than 20 reads. The initial exome analysis was negative; sequencing data was reanalyzed after this manuscript was deposited as a preprint in medRxiv (Redfield et al. 2023). Reanalysis revealed compound heterozygous variants in *PKHD1L1*, with the missense variant c.1813G>A, p,(Gly605Arg) inherited from the father, and a frameshift variant c.8452_8468del, p.(Leu2818TyrfsTer5) from the mother. The p.(Gly605Arg) missense variant was predicted to affect splicing by multiple tools, including dbNSFP (ada_score of 0.9998, and rf_score of 0.893), and SpliceAI (delta score of Donor Gain: 0.23). Interestingly, the same heterozygous missense variant c.1813G>A, p,(Gly605Arg) was found in another genetically undiagnosed proband with congenital bilateral severe SNHL from the Henan cohorts. In addition, the frameshift variant c.8452_8468del, p,(Leu2818TyrfsTer5) was also found in a genetically diagnosed proband (heterozygous c.8452_8468del; *MYO7A* c.689C>T, p.(Ala230Val), a known *MYO7A*pathogenic variant, (https://www.ncbi.nlm.nih.gov/clinvar/variation/178993/) (Di Leva et al. 2006; Kaneko et al. 2017; Lezirovitz and Mingroni-Netto 2022) with congenital bilateral SNHL from the Henan cohorts.

### Investigating the conservation of the mutated residue positions throughout evolution

All missense variants do not appear to cluster in any particular region of the *Hs* PKHD1L1 that was used for alignment and further analysis (Supplementary Fig. S1). In comparing the longest PKHD1L1 sequences among 10 different orthologs, we uncovered an overall amino acid sequence identity of 79.2 % (Supplementary Fig. S3). Notably, *Mm* and *Hs* PKHD1L1 share 81.8% of amino acid sequence identity (when comparing for identical sites excluding the signal peptides), while *Hs* and *Mm* orthologs of IPT1 and IPT6 shows 82.9% and 77.8%, respectively, suggesting high protein sequence conservation between the two species. Although some previous studies report protein sequence alignments and predictions of PKHD1L1 IPT domains (Hogan et al. 2003), an in-depth analysis was necessary to more accurately predict the signal peptide cleavage sites, as well as the starting and ending residues for each IPT domain. The location of the native Gly126, Gly1314, and His2479 residues, where the reported missense variants were detected, are highly conserved across a diverse set of the 10 different PKHD1L1 orthologues analyzed (Fig. 2d-e, Fig. 4a, and Supplementary Fig. S3 and Fig. S4).

### AlphaFold2 modeling of PKHD1L1 substitutions

PKHD1L1 has 14 predicted IPT extracellular-domain repeats of similar fold but with non-identical protein sequences and labeled as IPT1 to IPT14 from its N-terminal to C-terminal end, and other key domain features (Fig. 2b). The AlphaFold2 model of WT *Hs* IPT1-2 and its mutant p.(Gly129Ser) shows no apparent differences between their predicted structures (Fig. 3a-b), likely because the small side chain carrying this residue is located on a loop region exposed to the solvent. More specifically, the p.(Gly129Ser) variant is located within the connecting loop between the β-strand 6-7 of IPT1, close to a potential disulphide bond formed by p.Cys100 and p.Cys86, also found in plexin-like domains (Fig. 3b). Changes of the polarity or the electrostatic potential of this loop by p.(Gly129Ser) might cause structural changes or altered loop dynamics in IPT1 (Krieger et al. 2005). We also generated AlphaFold2 models for *Hs* IPT5-6 consistent with the expected IPT plexin-like folding for this structure (Fig. 3c-e). According to this structural model, the *Hs* p.(Gly1314Val) mutation is also located at the connecting loop between the β-strand 6-7 of IPT6 (Fig. 3c-e). Furthermore, the p.(Gly1314Val) variant is located within the connecting area between IPT5 and IPT6, and the AlphaFold2 model suggests a structural change for this specific fragment (Fig. 3d-e).

**Fig. 3.**
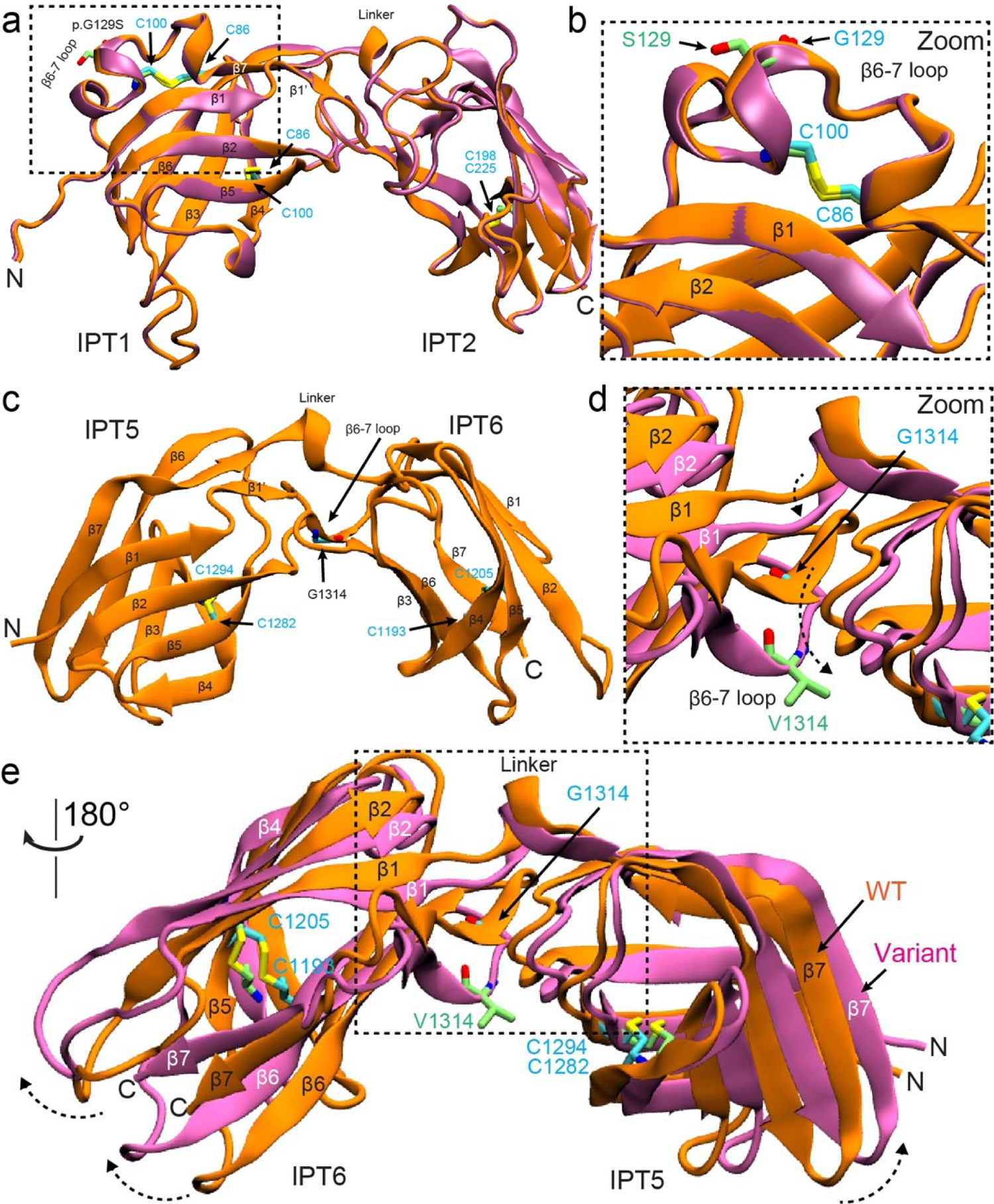
Alphafold2 modelling of PKHD1L1 protein fragments carrying Gly120Ser and Gly1314Val mutations. (a) Superposed AlphaFold2 models of both native *Hs* IPT1-2 (orange) and *Hs* IPT1-2 p.(Gly120Ser) variant (mauve) are shown. (b) Higher magnification image of the mutated site. p.S129 in lime and p.G129 in cyan. No apparent structural changes are predicted by AlphaFold2. (c) Structural model of *Hs* IPT5-6 showing the native Gly1314 position. (d) Superposed *Hs* IPT5-6 (orange) and *Hs* IPT5-6 p.(Gly1314Val) (mauve) structures showing a structural change predicted by AlphaFold2 as a result of p.(Gly1314Val) substitution. β-strands and loops do not overlap, with a dashed black arrow reporting the loop shift. (e) Higher magnification image of the mutated site showing the conformation change of β-strands and loops. p.V1314 (lime) cause steric hindrance in the area inducing an expanded conformation to the variant structure in mauve. See dashed arrows.

In a previous study, authors used protein sequence analysis of PKHD1, PKHD1L1, and TMEM2 reporting that PKHD1 and PKHD1L1 share two regions of significant sequence homology with TMEM2 (Hogan et al. 2003). AlphaFold2 modelling of the p.(His2479Gln) variant and surrounding PKHD1L1 region, revealed a high structural homology with a *Hs* TMEM2 protein (Fig. 4). We identified that this region features a conserved p.His2479 residue (throughout 10 different PKHD1L1 orthologs, Supplementary Fig. S4) (p.His552 in *Hs* TMEM2, Fig. 4a; Protein Data Bank (PDB): 8C6I (Niu et al. 2023)) reported to form a nickel-finger binding site, which might mediate catalytic functions in TMEM2. Disruption of this site in PKHD1L1 and TMEM2 might impair cation binding (Fig. 4a-f and Supplementary Fig. S3 and S4) and suggests a potential deleterious effect for this variant on protein structure and function. This locus is predicted neutral in terms of tolerance to missense substitution (Supplementary Fig. S2).

**Fig. 4.**
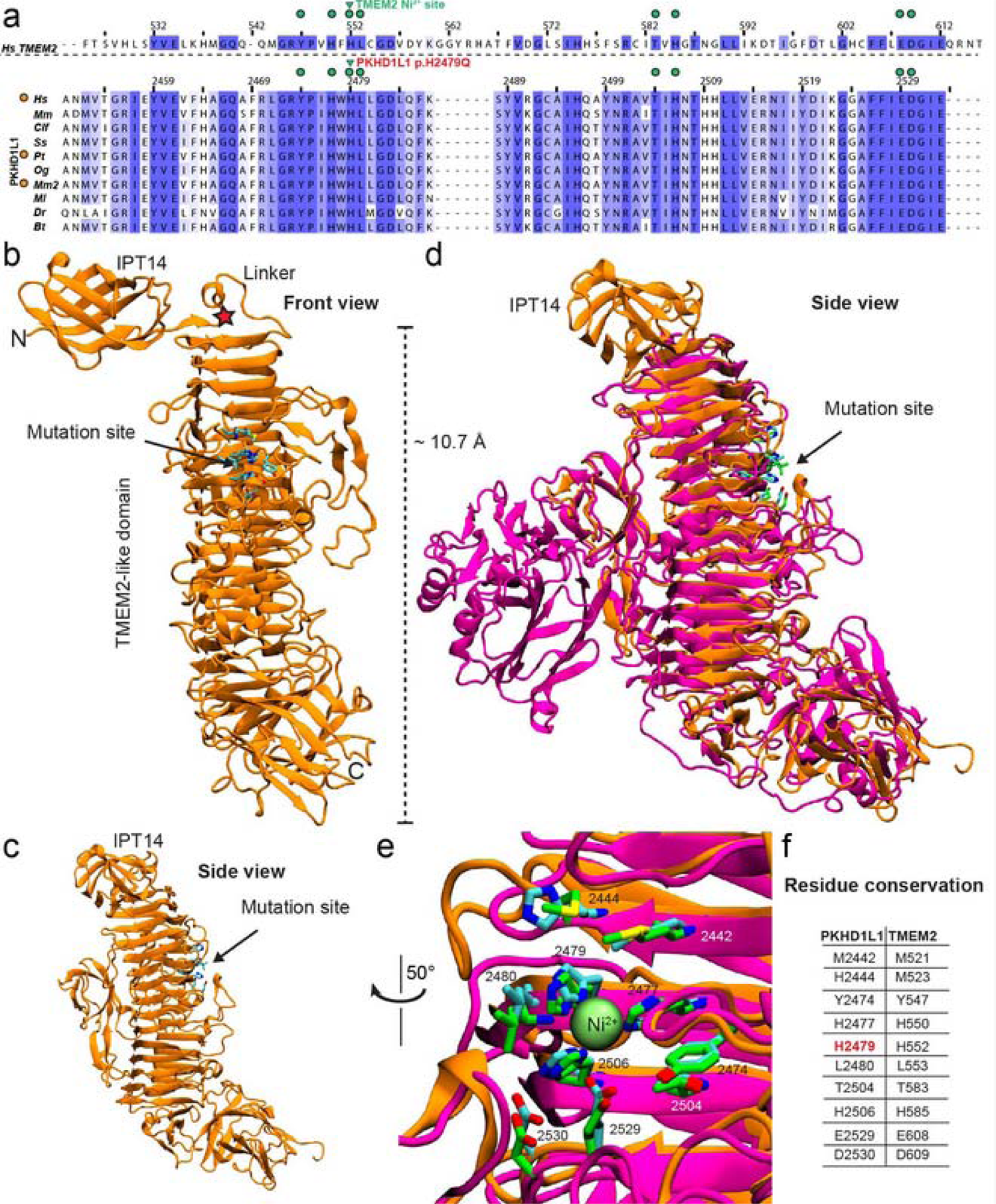
PKHD1L1 structural modelling of the protein fragment containing the p.(His2479Gln) variant. Based on AlphaFold2 predictions, this fragment of PKHD1L1 shares a common fold with the TMEM2 protein within the region carrying the p.(His2479Gln) variant. (a) Protein sequence alignment of a protein segment of *Hs* TMEM2 against 10 different PKHD1L1 orthologs (See Supplementary Table S1 for details of the selected species, Supplementary Fig. S4 for sequence alignment of this specific fragment, and Supplementary Fig. S3 for full PKHDL1 sequence alignment). Residue numbering for TMEM2 as in PDB: 8C6I (Niu et al. 2023), while residue numbering for *Hs* PKHD1L1 as in NCBI accession code NP_803875.2 with the signal peptide included (Supplementary Table S1, 26 residues are suggested according to protein sequence alignment, see Methods). Green triangles point to the location of the *Hs* p.(His2479Gln) variant, orange circles (*left*) indicate 100% amino acid sequence identity for this PKHD1L1 fragment between the *Hs*, *Pt*, and *Mm2* species (See supplementary Table S1 for details about the selected orthologs). Green circles represent depicted residues in panels b-e. The alignment was color-coded for sequence similarity (35% threshold) using Jalview. White-colored residues show the lowest similarity and dark blue report the highest (see Methods). PKHD1L1 orthologs were chosen based on sequence availability and taxonomical diversity. (b) The simulated protein structure covering the protein fragment highlighted by purple arrow-headed line in Fig. 2b. Front view of the structure showing IPT14 linked to the PKHD1L1 TMEM2-like domain. The red star points to the linker connection. Residues at the mutation site are depicted as cyan sticks. (c) Side view from *panel a* showing a clear view of the stacked β-strand motifs. (d) Superposed structural protein alignment between WT *Hs* PKHD1L1 TMEM2-like domain model (orange) with the X-ray crystal structure of *Hs* TMEM2 protein (PDB: 8C6I, magenta). Residues at the native TMEM2-histidine finger site are depicted as green sticks. (e) Higher magnification image of the potential conserved histidine-finger site between PKHD1L1 (orange) and TMEM2 (magenta) protein fragments and the Ni^2+^ ion shown as lime sphere. (f) Displayed residues between both proteins surrounding the Ni^2+^-ion site highlighted in *panel a* in green circles.

### Functional testing of the p.(Gly129Ser) and p.(Gly1314Val) substitutions

Next, we expressed and purified the recombinant WT *Mm* PKHD1L1 IPT1-3 and IPT5-6 protein fragments as well as the respective IPT1-3 p.(Gly129Ser) and IPT5-6 p.(Gly1314Val) mutant protein fragments using SEC (Supplementary Fig. S5). These protein constructs represent key regions of the complete extracellular domain of PKHD1L1 where these mutations might locally affect the structural assembly of the protein. The thermodynamic and folding stabilities were measured using NanoDSF to assess the protein stabilities in solution for WT PKHD1L1 protein fragments and compared to fragments carrying missense mutations (Fig. 5 and Supplementary Fig. S5). For WT IPT1-3, the T_onset_ (melting temperature at which unfolding begins) was measured at a maximum of 58 °C, while the T_onset_ for IPT1-3 p.(Gly129Ser) variant was 52 °C (a 6 °C decrease, Fig. 5 and Supplementary Fig. S5). In addition, there was a decrease on the T_m_ (melting temperature or point at which 50% of the protein is unfolded) of ∼ 4 °C comparing different thermal transition points between WT and the IPT1-3 p.(Gly129Ser) variant (Fig. 5a). These measurements strongly suggest that the p.(Gly129Ser) variant affects PKHD1L1 protein stability within this region.

**Fig. 5.**
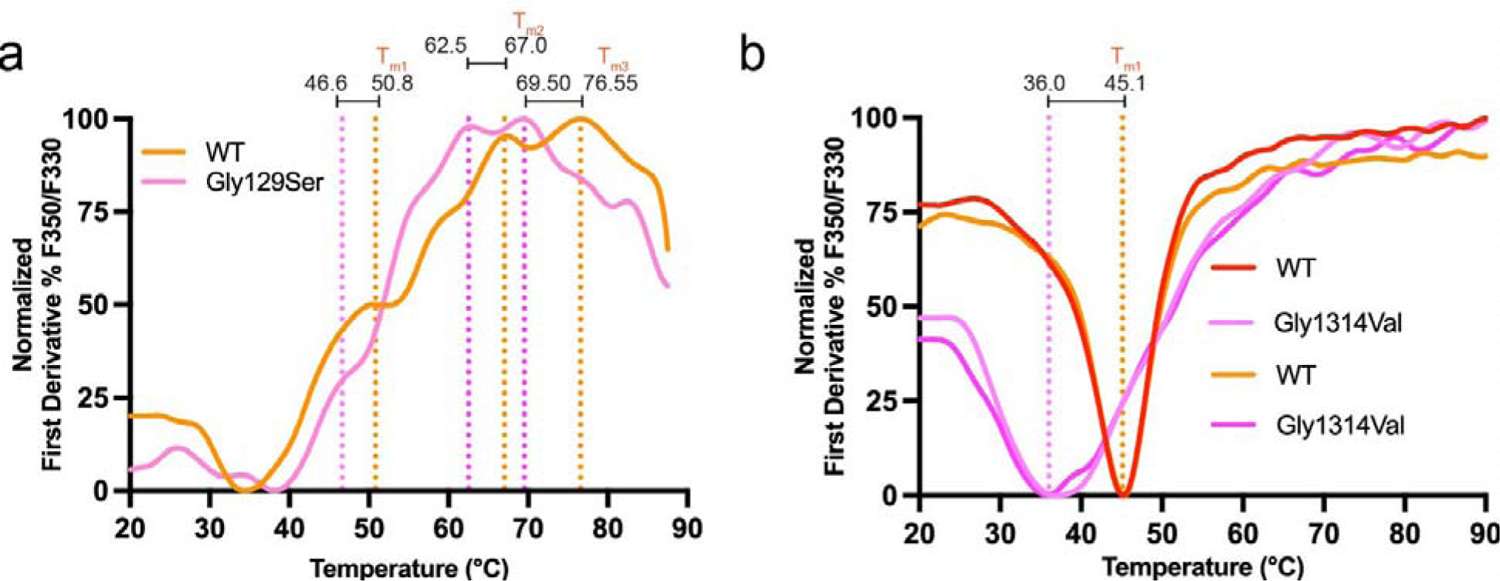
Thermodynamic and folding stability evaluation of two missense variants using NanoDSF. (a) NanoDSF melting temperatures for WT *Mm* IPT1-3 (orange) and *Mm* IPT1-3 p.(Gly129Ser) variant (pink). Measurements show at least three T_m_ peaks (orange dotted line) for the WT IPT1-3, likely because the protein fragment includes multiple IPT domains that unfold sequentially. Measured T_m_ values are shifted to the left (pink dotted line) showing a decrease of the thermal folding stability. Temperatures are labeled for each T_m_ transition point. (b) Results for WT *Mm* IPT5-6 and *Mm* IPT5-6 p.(Gly1314Val) showing a reduced thermal stability (2 replicates, see Methods section). Traces correspond to the normalized first derivate of the fluorescence ratio showing the inflection point of the fluorescence ratio, which corresponds to the melting temperature of the sample. T_onset_ values and protein purification experiments are shown in Supplementary Fig. S5.

Similarly, NanoDSF measurements for WT *Mm* PKHD1L1 IPT5-6 and *Mm* IPT5-6 p.(Gly1314Val) showed a T_onset_ of 35.48 °C and 28.56 °C, respectively (∼ 7 °C decrease, Fig. 5 and Supplementary Fig. S5). Additionally, WT *Mm* PKHD1L1 IPT5-6 showed a melting temperature T_m_ of 45.10 °C, while the mutant IPT5-6 p.(Gly1314Val) displayed a decrease on this T_m_ to 36.6 °C (decreasing the temperature ∼ 8.6 °C) (Fig. 5b). These results confirm that both IPT1-3 p.(Gly129Ser) and IPT5-6 p.(Gly1314Val) variants indeed significantly decrease the thermal and folding stabilities of these PKHD1L1 protein fragments.

### Splicing evaluation of p.(Gly605Arg)

Based on *in silico* evaluation (Table 1), the missense variant p.(Gly605Arg) was predicted to affect splicing given that it occurs at the 3’ exon boundary of exon 17 adjacent to the 5’ intronic splice donor site of intron 17 (Fig. 6). RNA studies of this variant indicated a functional effect on splicing, leading to an in-frame deletion of 48 amino acids (r.1670_1813del, p.Val557_Arg604del; Fig. 6).

**Fig. 6.**
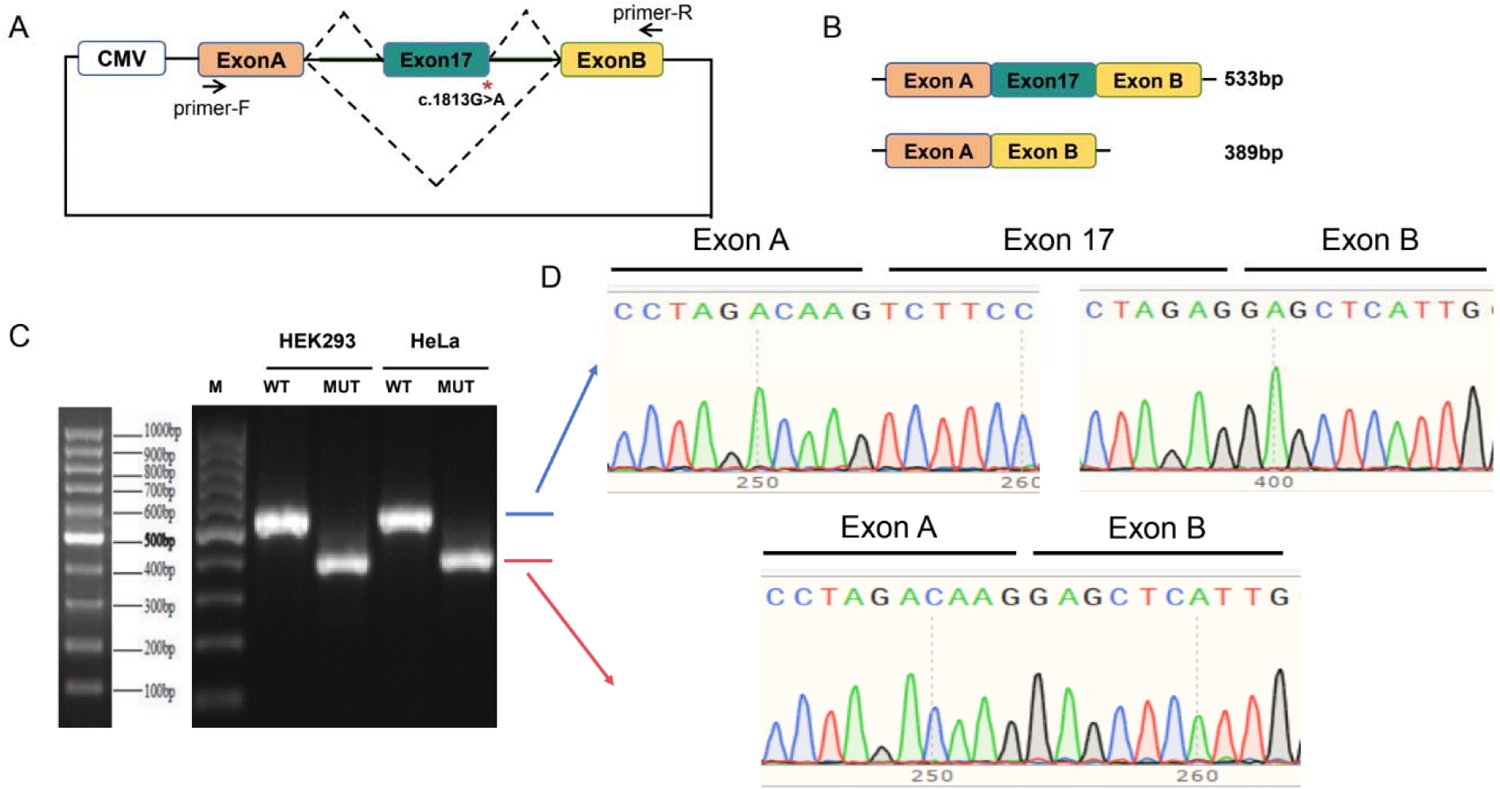
Minigene splicing assay for evaluation of the functional effect of p.(Gly605Arg) on splicing. (a) Schematic demonstrating designed minigene assay including CMV promotor, mutation location, and primers. (b) Schematic showing calculated size of fragment with exon 17 included (533 bp) or excluded (389 bp). (c) and (d) RT-PCR result from HEK293 and HeLa cells transfected with both WT and mutant plasmids showing different fragment lengths as well as sequence on chromatogram demonstrating lack of incorporation of exon 17 in cells transfected with the mutant plasmid, leading to in-frame deletion of 48 amino acids (p.Val557_Arg604del) using the NCBI NP_803875.2 as a reference sequence.

## Discussion

A majority of congenital SNHL is attributable to a genetic etiology, and clinical genetic testing for known SNHL genes is an established standard of care in the diagnostic evaluation of bilateral SNHL in pediatric patients (Shearer and Smith 2015; Smith et al. 2005). To date, there are over 120 known genetic causes of nonsyndromic hearing loss, and gene panel tests are recommended to facilitate accurate and timely genetic diagnosis of SNHL (https://hereditaryhearingloss.org). However, despite advances in genetic testing for SNHL, the diagnostic yield for SNHL ranges from 22.5% to 55.7% with an average of ∼ 40% (Downie et al. 2020; Perry et al. 2023; Rouse et al. 2022; Sloan-Heggen et al. 2016). The identification of novel hearing loss genes is critical to improving diagnostic rates, thus impacting care and management for individuals with SNHL.

In mice, PKHD1L1 is predominantly expressed in the OHC stereocilia by P0 to P12 with a basal-to-apical (decreasing) expression gradient and is a major component of the stereocilia surface coat (Wu et al. 2019). *Pkhd1l1*-deficient mice lack the surface coat at the stereocilia tips and exhibit progressive SNHL by ABR and DPOAE measurements starting as early as 3 weeks. Although its function remains largely undetermined, the two functional hypotheses of PKHD1L1 expression at the stereocilia include that it may be required for the correct localization of other stereociliary proteins, or it plays a role during the development of attachment crowns at the stereocilia to secure the tectorial membrane to the bundle. An immature attachment could manifest as a persisting relaxed tectorial membrane coupling (Wu et al. 2019). Furthermore, it is unknown whether PKHD1L1 is the only component of the stereocilia coat. Recently, *pkhd1l1* was shown to play a critical role in regulating hearing in zebrafish (Makrogkikas et al. 2023). *pkhdl1l* has a ubiquitous expression pattern and is sustained for most of embryonic development (Makrogkikas et al. 2023). Through depletion of the two paralogous genes (*pkhd1l1a* and *pkhd1l1b*), double mutant zebrafish exhibited significant hearing loss from the larval stage (6 days post fertilization) which differs compared to progressive hearing loss in the mouse (Wu et al. 2019).

Although presenting congenitally in the majority of patients, the degree of hearing impairment in the patients we present is fairly broad: The proband in family 1, with p.[(Gly129Ser)];p.[(Gly1314Val)] compound heterozygous variants, was diagnosed with congenital hearing impairment that remains mild to moderate at the age of 11-15 years; the proband in family 2, with a homozygous p.(Arg3381Ter) variant, already showed moderate to severe SNHL at the age of 6-10 years; and the proband in family 3, at the age of 11-15 years, showed severe hearing impairment attributed to the homozygous p.(His2479Gln) variant. However, in case of the proband in family 3 with the PKHD1L1 .p(His2479Gln) variant, it is possible that notwithstanding neutral predictions by different software packages (Table 1, and Supplementary Fig. S2), the detected MYO7A p.Leu375Val missense variant is pathogenic (Supplementary Table S2) and the *PKHD1L1* variant is an incidental finding. Or, both *MYO7A* and *PKHD1L1* variants may contribute to the more severe hearing loss of this individual. However, AlphaFold2 modelling showed that the MYO7A p.Leu375Val variant might not induce structural changes in MYO7A (Supplementary Fig. S6). The proband in family 4 with p.[(Gly605Arg);p.(Leu2818TyrfsTer5)] compound heterozygous variants had a moderate hearing loss at the age of 6-10 years.

While we have identified individuals in four families with variants in *PKHD1L1*, this study highlights the necessity for an extended case series with longitudinal audiological follow up and functional studies to assess variant effects of patient-specific perturbations on development, maturation, and function of the auditory system, as well as explore the potential of accelerated effects of age, noise, or trauma on progression of hearing loss, which remain as current major limitations. Interestingly, the *PKHD1L1* gene has been suggested to be associated with adult-onset hearing loss (Lewis et al. 2023). Since the studied variants are also located in different residue positions in the PKHD1L1 protein sequence, the broad range of hearing impairment from these patients might suggest that these variants differentially impact the protein expression, folding, and/or the stability and function of PKHD1L1. Therefore, we cannot exclude an environmental component that may account for variability.

We also investigated the conservation of the mutated residue positions throughout evolution. Multiple sequence alignments of the complete PKHD1L1 amino acid sequence from 10 different orthologs were analysed and found to be highly conserved. This suggests that these native residues might be critical to protein folding and assembly. Therefore, variants in these positions might disrupt protein function and potentially cause hearing impairment *in vivo*.

Because the p.(Arg3381Ter) introduces a stop codon that would be predicted to be targeted by nonsense mediated decay by the 50-nucleotide rule (Frischmeyer and Dietz 1999), it is anticipated that this would result in the lack of protein or low yield of truncated protein expression without the transmembrane domain, key for the proper insertion into the cell membrane. This is likely to impair the proper folding, trafficking, and insertion of PKHD1L1 in the stereocilia-plasma membrane, or even result in secretion of PKHD1L1 extracellular fragments that could progressively affect hearing function. Interestingly, secreted versions of extracellular PKHD1L1 have been found in supernatant solution from platelet cells (Maynard et al. 2007) and their soluble concentrations could be modulated by protease inhibitors (Fong et al. 2011), suggesting potential cleavage sites in *Hs* PKHD1L1. However, the role of these potentially cleaved extracellular PKHD1L1 protein fragments remains unknown.

To further predict how these PKHD1L1 mutant variants might affect the PKHD1L1 protein at the structural level, we modelled the structures of the individual domains carrying reported variants (Fig. 3a-e and Fig. 4b-e). The p.(Gly129Ser) substitution in IPT1 was not predicted to exert an apparent structural difference. We speculate that instead changes of the polarity or the electrostatic potential of the β-strand linker loop by p.(Gly129Ser) might alter loop dynamics in IPT1. Interestingly, both glycine substitutions p.(Gly129Ser) and p.(Gly1314Val) are located within the same connecting loop between β-strand 6-7 in IPT1 and IPT6, respectively. While the AlphaFold2 model of the p.(Gly129Ser) mutant shows no apparent structural changes in the predicted structure (Fig. 3a-b), the AlphaFold2 model of p.(Gly1314Val) shows a conformation change, likely due to steric hindrance on the structure (Fig. 3c-e). Finally, we also mapped the location of the p.(His2479Gln) using structural modeling by AlphaFold2 (Fig. 4, Supplementary Fig. S4). Our results indicate that the His2479 position (among 10 different PKHD1L1 orthologs) is 100% conserved with a *Hs* TMEM2 protein (PDB: 8C6I), a regulator of the hyaluronan metabolism (Fig. 4a and 4e-f) (Sato et al. 2023). Interestingly, experiments suggest that TMEM2 activity is calcium-dependent (Yamamoto et al. 2017) and TMEM2 has been previously studied for its structural similarities with the *CEMIP* deafness gene candidate (Yoshida et al. 2013).

Our NanoDSF thermal folding analysis showed that both T_onset_ and T_m_ values decreased in the presence of the p.(Gly129Ser) and p.(Gly1314Val) variants. The T_m_ measurements using NanoDSF showed p.(Gly129Ser), located in a loop, decreases the stability of IPT1 and further showed how this variant propagates its destabilizing effects to the neighboring IPT2 and IPT3 (Fig. 2b, Fig. 5, and Supplementary Fig. S5). Likely, the p.(Gly1314Val) variant also alters the stability of the loop and the chemical environment in the IPT5-IPT6 connection, since the measured folding stability showed an 8.6 °C decrease of unfolding temperature between WT IPT5-6 fragment and the p.(Gly1314Val) variant (Fig. 5). This is the first study showing strong evidence to support how missense variants locally affect the structural folding and stability of PKHD1L1 fragments *in vitro*. Given the high conservation rate of 81.8% in amino acid sequence identity between the *Hs* and *Mm* PKHD1L1 (excluding the 20 amino acid long signal peptide according to SMART, Supplementary Fig. S3), and the 100% conservation of the mutated sites across the species analyzed, we believe our findings using *Mm* PKHD1L1 protein fragments can be directly applied to *Hs* PKHD1L1. Future functional studies involving highly conserved full-length PKHD1L1 orthologs and their protein purification would allow for better understanding of various effects such variants might have on the stability of the entire PKHD1L1 extracellular domain, its protein expression and proper localization, which might be linked to the progression and hearing loss severity. Furthermore, studies focused on uncovering the influence of mutations on the structure of the complete PKHD1L1 extracellular domain will help to better understand the role of PKHD1L1 in hearing function and beyond, since the PKHD1L1 has been suggested as a tumor suppressor (Yang et al. 2023) and a human cancer biomarker (Kafita et al. 2023; Wang et al. 2023; Zheng et al. 2019).

As we have shown that single point mutations have a detrimental effect on protein folding and stability in protein fragments using NanoDSF, the deletion of longer protein motifs in PKHD1L1 extracellular domain might contribute to a more detrimental effect. In this case, the *in vitro* evidence presented in this study for the PKHD1L1 p.(Gly605Arg) missense variant found in the proband of family 4 (Fig. 1, inherited from the father), strongly supports that this post-transcriptional splicing modification leads to an in-frame deletion of 48 amino acids (p.Val557_Arg604) (Fig. 6). This could explain the more significant hearing deficit caused by the p.(Gly605Arg) splicing mutation along with the frameshift variant c.8452_8468del, p.(Leu2818TyrfsTer5) (inherited from the mother) in the same individual, compared to the milder hearing loss phenotype presented in the proband in family 1 (compound heterozygous p.[(Gly129Ser)];p.[(Gly1314Val)]).

Additional syndromic involvement was excluded in all four probands. However, in addition to hearing impairment, disruption of PKHD1L1 has also been associated with increased susceptibility to pentylenetetrazol-induced seizures in mice indicating a possible role in maintenance of neuronal excitability in the central nervous system (Yu et al. 2023). It is currently unknown whether defects in *PKHD1L1* might cause sensory auditory seizures. However, PKHD1L1 is expressed in the hippocampus and cerebral cortex in adult WT mice. Knockdown of *PKHD1L1* using *PKHD1L1*-shRNA or *PKHD1L1*-shRNA-AAV increased susceptibility of seizures as indicated by increased epileptiform bursting activity in cultured hippocampal neurons and pentylenetetrazol-induced seizures of mice following knockdown, suggesting a role for PKHD1L1 in the maintenance of normal excitation-inhibition balance (Yu et al. 2023). Knockdown of *PKHD1L1* led to the downregulation of both expression and function of the KCC2 membrane protein, which may explain the increased susceptibility to seizures (Yu et al. 2023). There is no evidence that mutations in *PKHD1L1* lead to seizures in humans, though an open question remains as to whether similar pleiotropic effects paralleling, for example, the various phenotypes caused by pathogenic variants in *TBC1D24* may also occur as more *PKHD1L1* patients are discovered (Mucha et al. 1993; Rehman et al. 2017).

## Conclusion

Here we provide data to support that mutations in *PKHD1L1* cause human nonsyndromic autosomal recessive congenital, mild-moderate to severe SNHL. We demonstrated that all reported missense variants point to highly conserved residues throughout evolution, suggesting that the native residues are key for protein folding and function, while variants in these sites locally affect the thermal-folding stability of PKHD1L1 fragments in solution. Inclusion of *PKHD1L1* as a hearing loss gene is supported by four families segregating plausible variants, *in vitro* functional data confirming their detrimental impact, as well as previously published mouse and zebrafish models demonstrating hearing loss. This study serves as a call to clinical laboratories to include careful screening of *PKHD1L1* biallelic variants in patients with a hearing loss ranging from mild-moderate to severe. Further research will be needed to determine the effect of age, noise, or trauma on the potential progression of PKHD1L1 linked hearing loss.

## Data Availability

All data produced in the present study are available upon reasonable request to the authors

## Statements and Declarations Acknowledgments

We thank the families for their participation. We thank Mahdiyeh Behnam for support with CNV analysis for the proband in Family 3.

## Author contributions

SER and PD contributed equally as shared first authors, MZ and HW contributed equally as second authors, and AAI, AES and BV contributed equally as shared last authors. AES, PD, and AAI conceived the study. PD, TM, and AAI designed methodology. PD and TM contributed software. MZ, PD, and AAI performed formal analyses that were validated by HG, MK, PD, TM, and AAI. MZ, HW, HK, MK, SN, MAK, PD, TM, AAI, and BV conducted the investigation process. HG, GS, MAK, HX, WL, AES, SER, and PD secured patient and experimental resources. MZ, HW, HG, GS, BV, SER, WL, AES, PD, and AAI performed data curation. BV, AES, PD, and AAI wrote the original draft. MZ, HK, MK, SN, HG, BV, SER, AES, PD, and AAI contributed to reviewing and editing. BV, AES, SER, PD, and AAI prepared figures. SN, AES, PD, and AAI provided supervision. BV, AES, and AAI oversaw project administration. All authors reviewed and approved of the manuscript.

## Compliance with ethical standards Conflict of interest

Go Hun Seo is an employee of 3Billion, Inc. The other authors have no conflicts to declare.

## Ethics approval

This study was approved by the institutional review boards of Boston Children’s Hospital (IRB P-00031494), University Medical Center Göttingen (No. 3/2/16), and the School of Biological Sciences, University of Punjab, Lahore, Pakistan (IRB No. 00005281).

## Consent to participate

Written informed consent was obtained from all participants.

## Consent to publish

Consent to publish was obtained from all participants.

