## Supplementary material for "*PKHD1L1*, A Gene Involved in the Stereocilia Coat, Causes Autosomal Recessive Nonsyndromic Hearing Loss": Suppl Fig. S1, Suppl Fig. S2, Suppl Fig. S3, Suppl Fig. S4, Suppl Fig. S5, Suppl Fig. S6, Suppl Table S1, Suppl Table S2

**Supplementary Fig. S1.** *PKHD1L1* transcript schematic

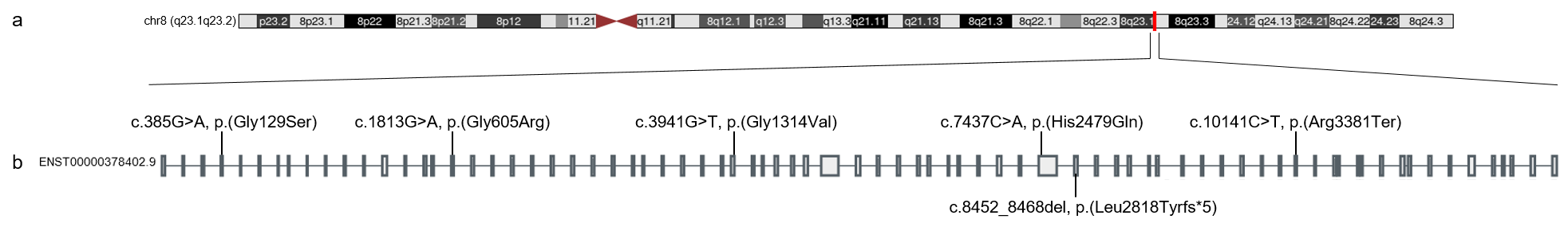

**Supplementary Fig. S1.** *PKHD1L1* transcript schematic

(a) *PKHD1L1* maps to human chromosome 8q23.1q23.2 (boxed in red).

(b) Overview of the human PKHD1L1 gene (NCBI code: NM_177531.6) from the GTEx Portal with annotated variant positions.

**Supplementary Fig. S2.** Predicted tolerance landscape of *PKHD1L1* missense variants

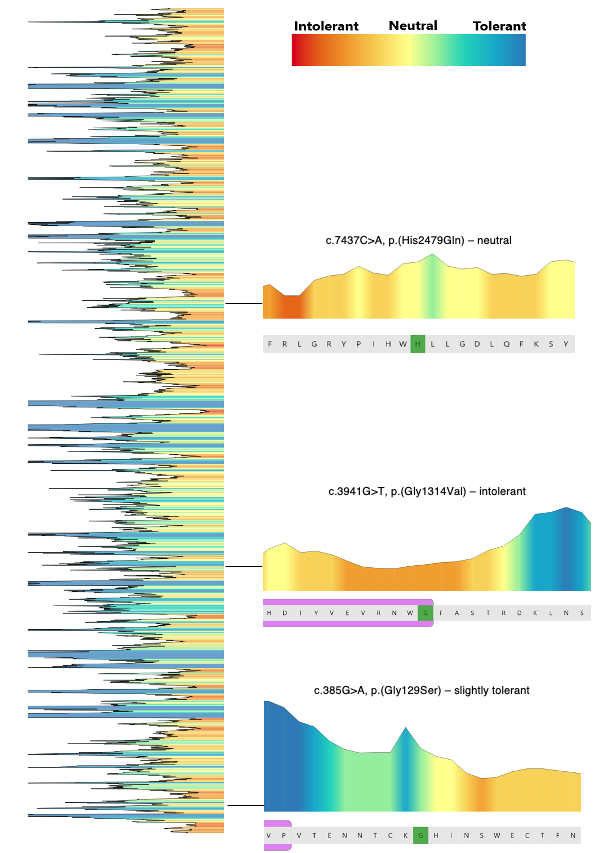

**Supplementary Fig. S2.** Predicted tolerance landscape of *PKHD1L1*

MetaDome web server was used to query each amino acid substitution in our *PKHD1L1* cohort to estimate tolerance/intolerance to substitution. The overview of the predicted tolerance landscape is shown on the left panel. Each of the three amino acid substitutions are shown with a zoomed in region with 10 flanking amino acids and residue of interest marked in green. Protein domains are denoted in magenta. A scale from red (most intolerant) to blue (most tolerant) to amino acid substitutions is shown at the top. Note: as the c.1813G>A p.(Gly605Arg) exhibited an aberrant splicing effect, this variant was not included in the predicted landscape analysis.

**Supplementary Fig. S3.** PKHD1L1 protein sequence alignment of the complete sequence of 10 different species

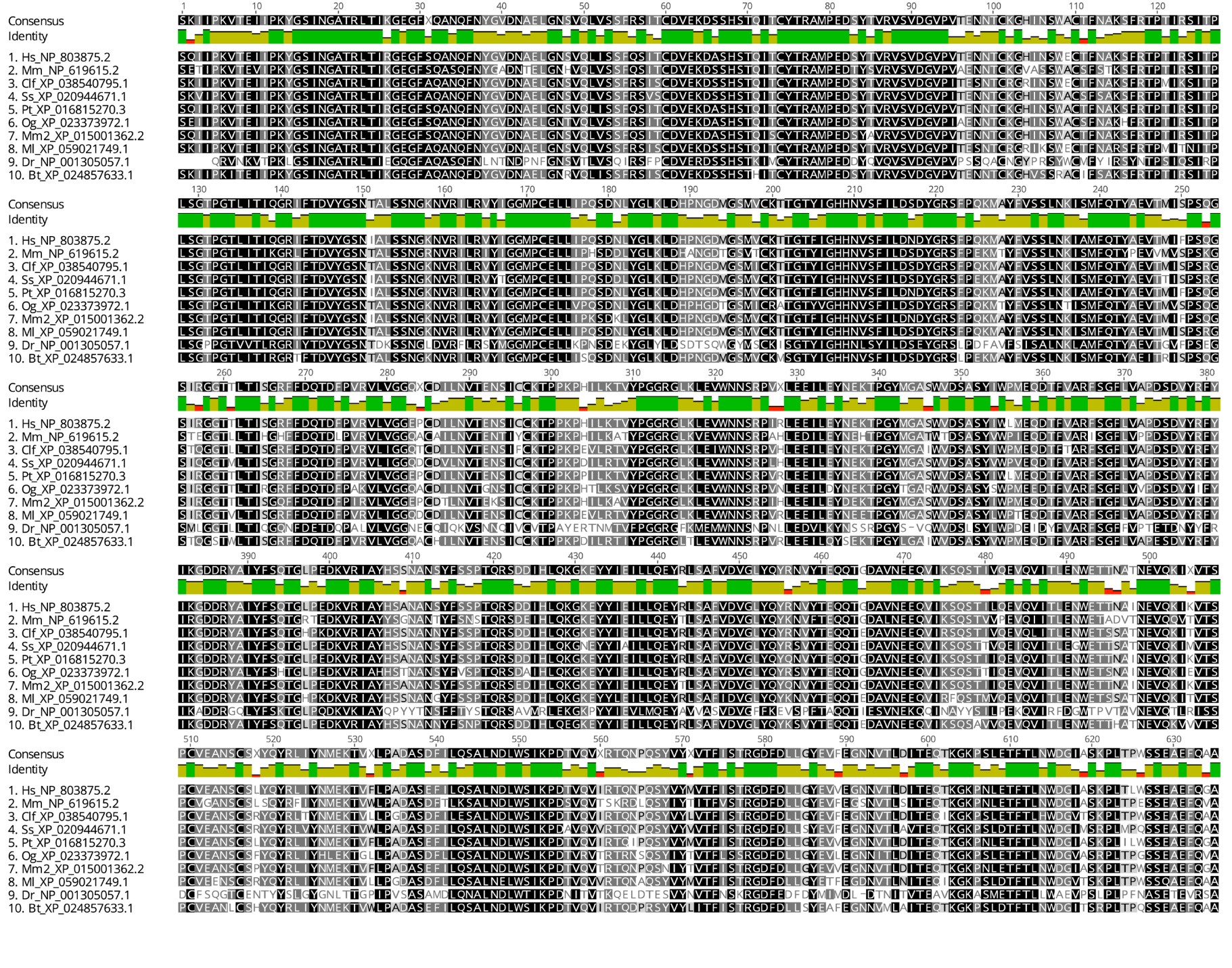

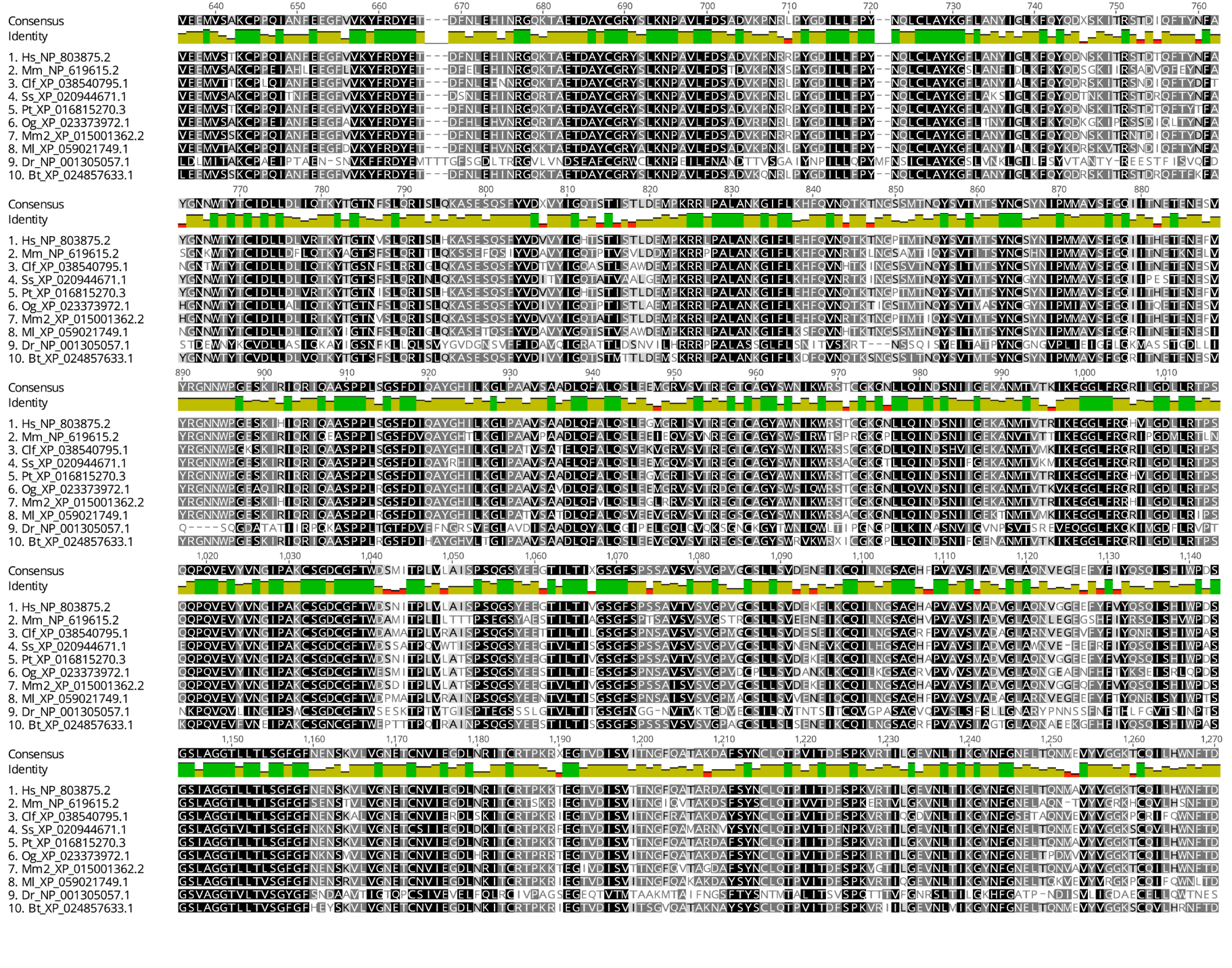

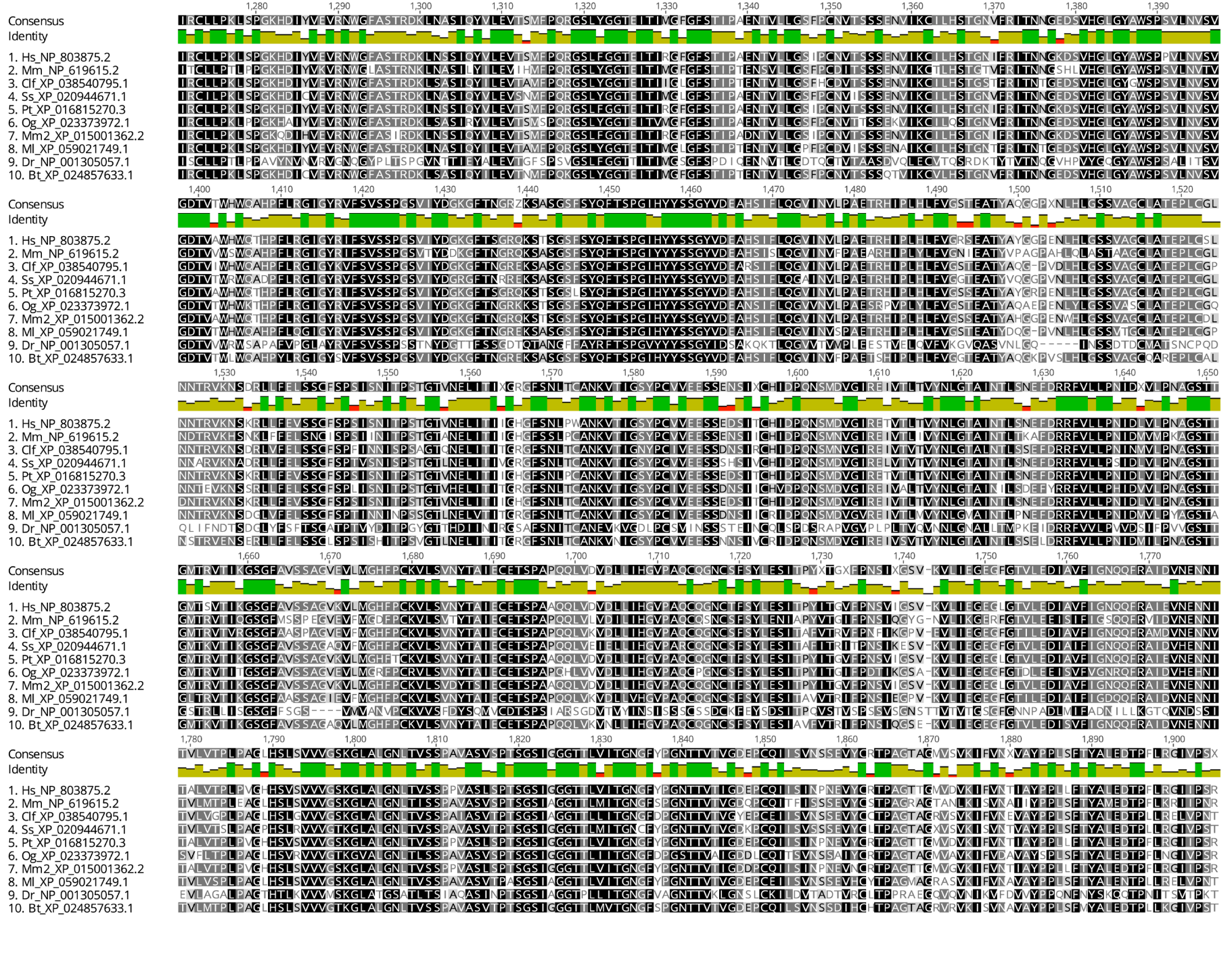

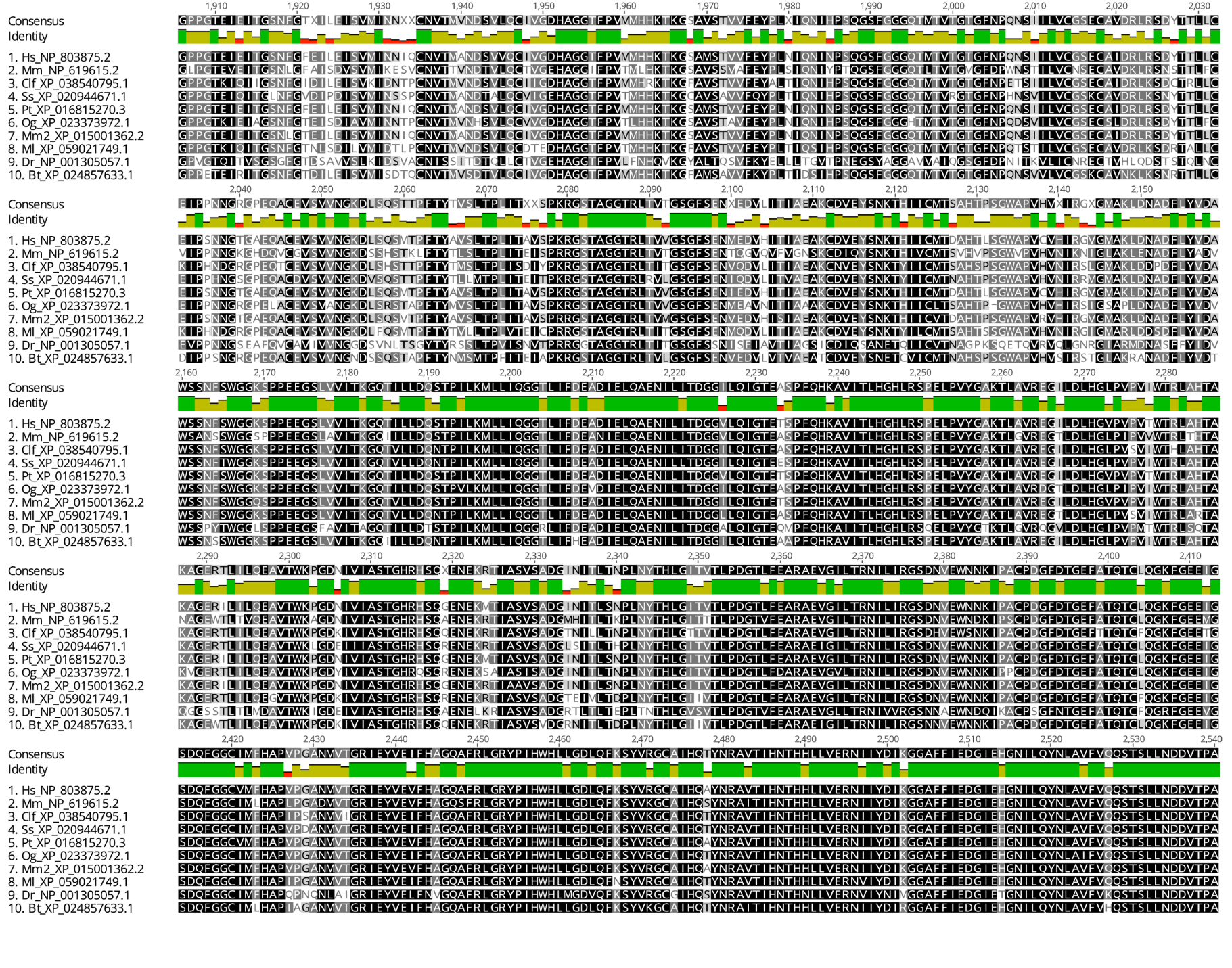

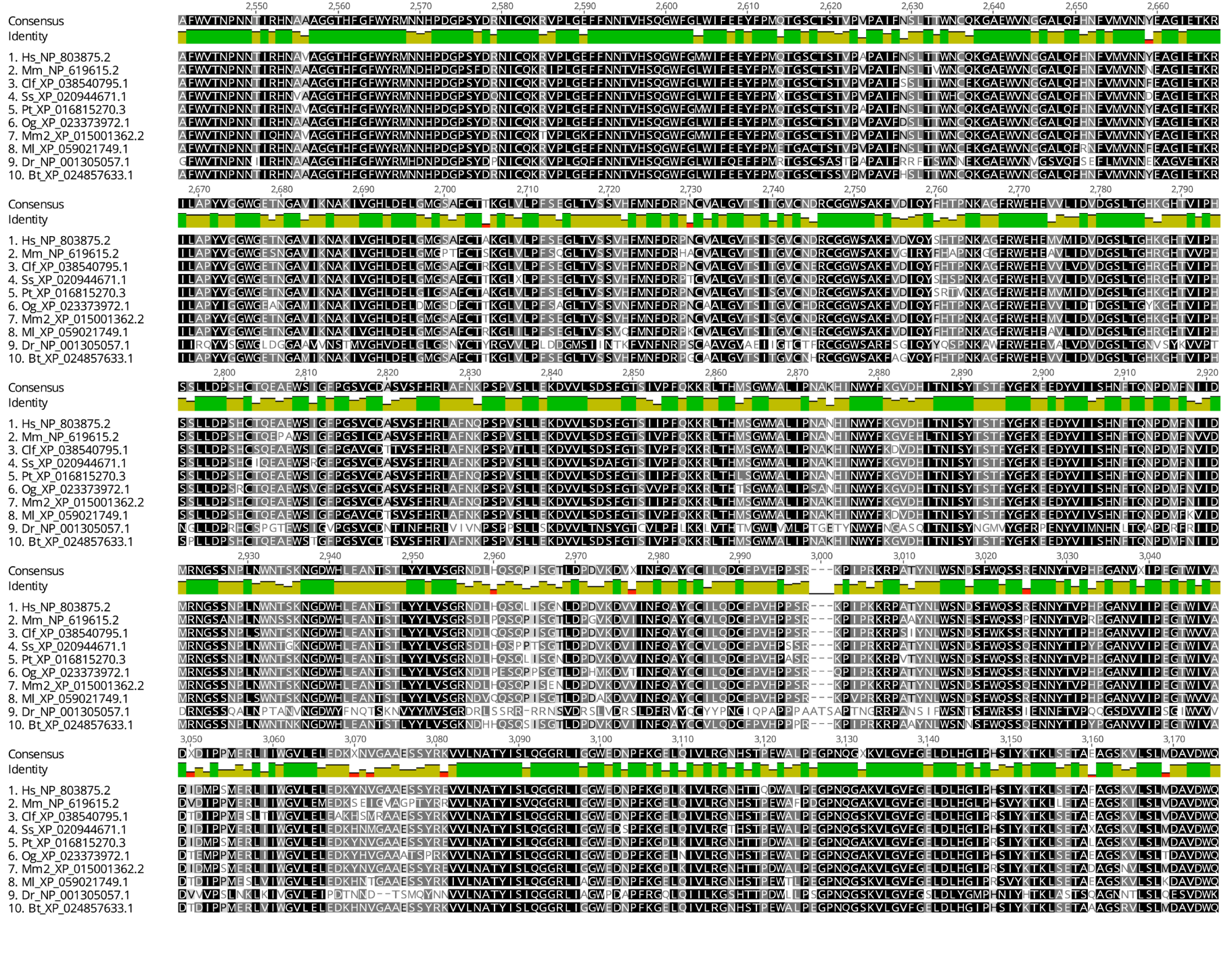

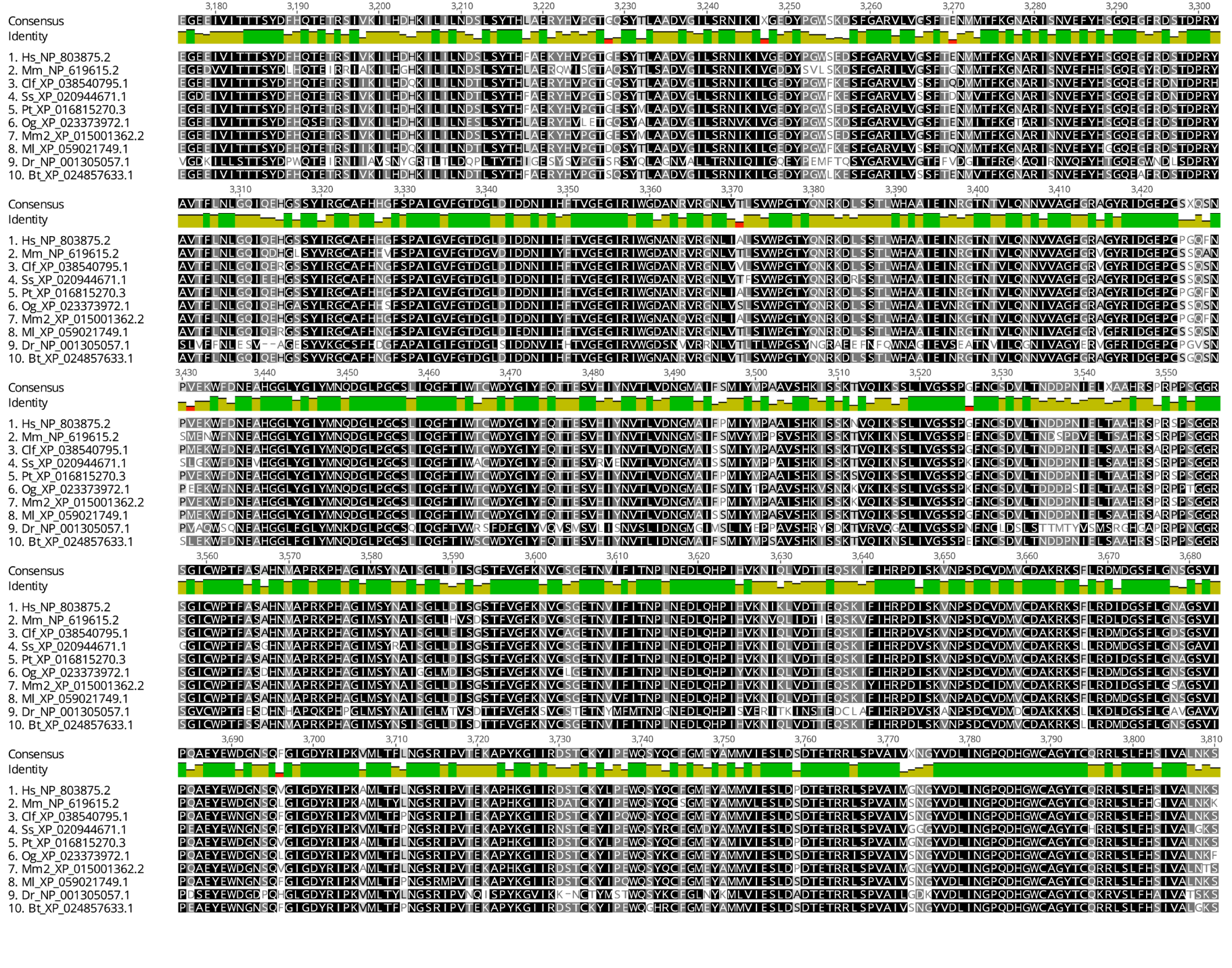

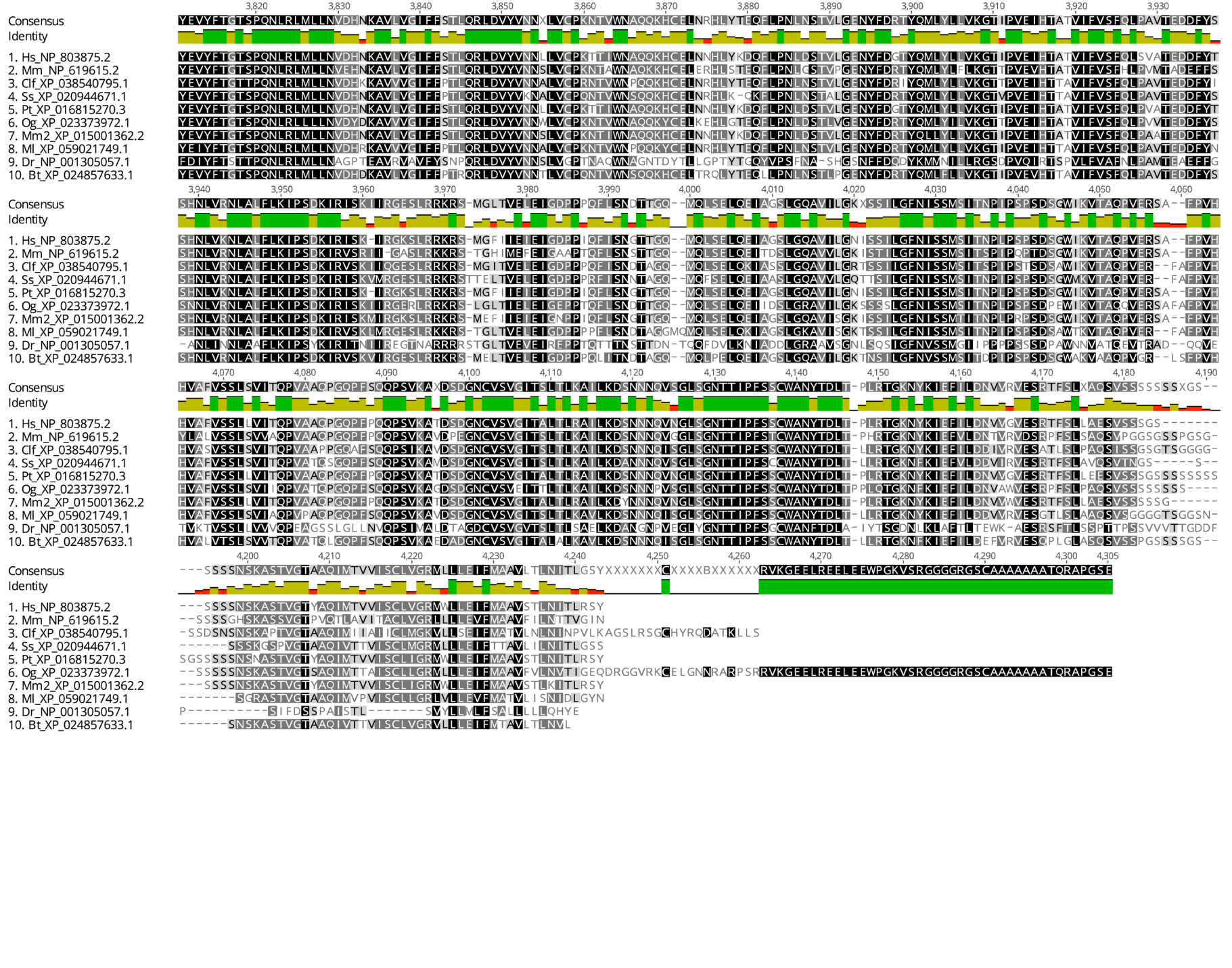

**Supplementary Fig. S3.** PKHD1L1 protein sequence alignment of the complete sequence of 10 different orthologs.

The complete PKHD1L1 sequence alignment across 10 species shows a high pairwise sequence identity of 79.2%. Notably, *Mm* and *Hs* PKHD1L1 share 81.8% of amino acid sequence identity these two orthologs. Protein sequences were extracted from the NCBI database. A consensus sequence is shown, and residue numbering exclude the signal peptide for each protein (20 amino acids for *Hs* PKHD1L1 based on SMART prediction (Letunic et al. 2021) and 26 on sequence alignment with the other PKHD1L1 orthologs). See Supplementary Table S1 for details about the species. Sequences were aligned with Geneious Prime and color-coded based on residue similarity. White-colored residues show the lowest similarity and black report the highest.

**Supplementary Fig. S4.** Protein sequence alignment of the region covering the PKHD1L1 TMEM2-like domain.

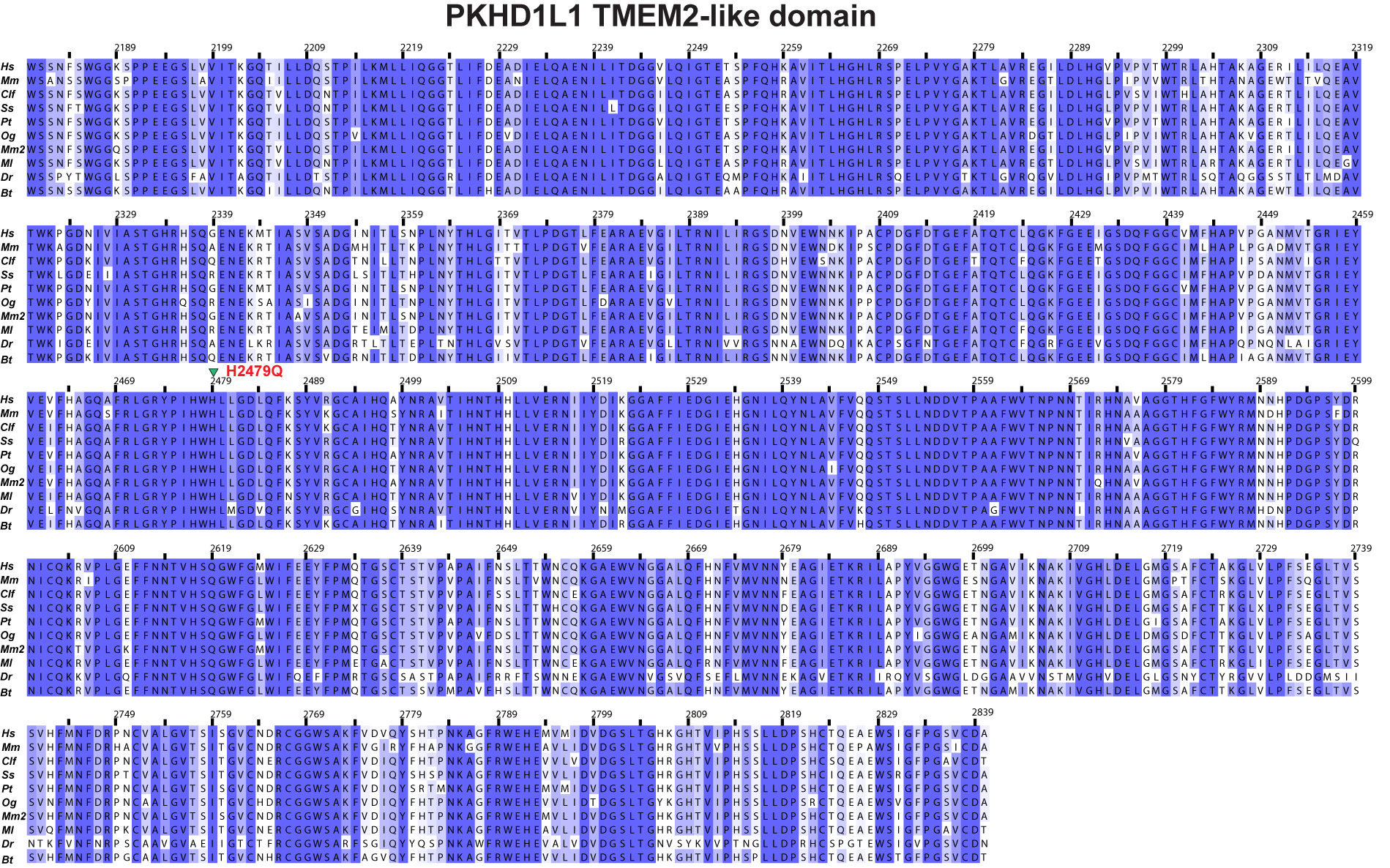

**Supplementary Fig. S4.** Protein sequence alignment of the region covering the PKHD1L1 TMEM2-like domain across 10 different species. The analyzed PKHD1L1 fragment sequence shows a high pairwise sequence identity of 88.3%.

The protein region presented in this figure is highlighted by a purple arrow-headed line in Figure 2b, and 10 different species of PKHD1L1 proteins were used. Alignment was color-coded for sequence similarity (35% threshold) using Jalview. White-colored residues report the lowest similarity and dark blue report the highest (see *Methods*). Species were chosen based on sequence availability and taxonomical diversity. Alignment results indicate that this fragment region shares 421 identical sites (63.7%). Residue numbering includes the signal peptide using *Hs* PKHD1L1 protein sequence as a reference (26 amino acids for *Hs* were predicted, see Supplementary Table S1 for NCBI accession numbers). The p.(His2974Gln) variant is highlighted in red and labeled with a green triangle.

**Supplementary Fig. S5.** PKHD1L1 fragment protein purification and NanoDSF experiments.

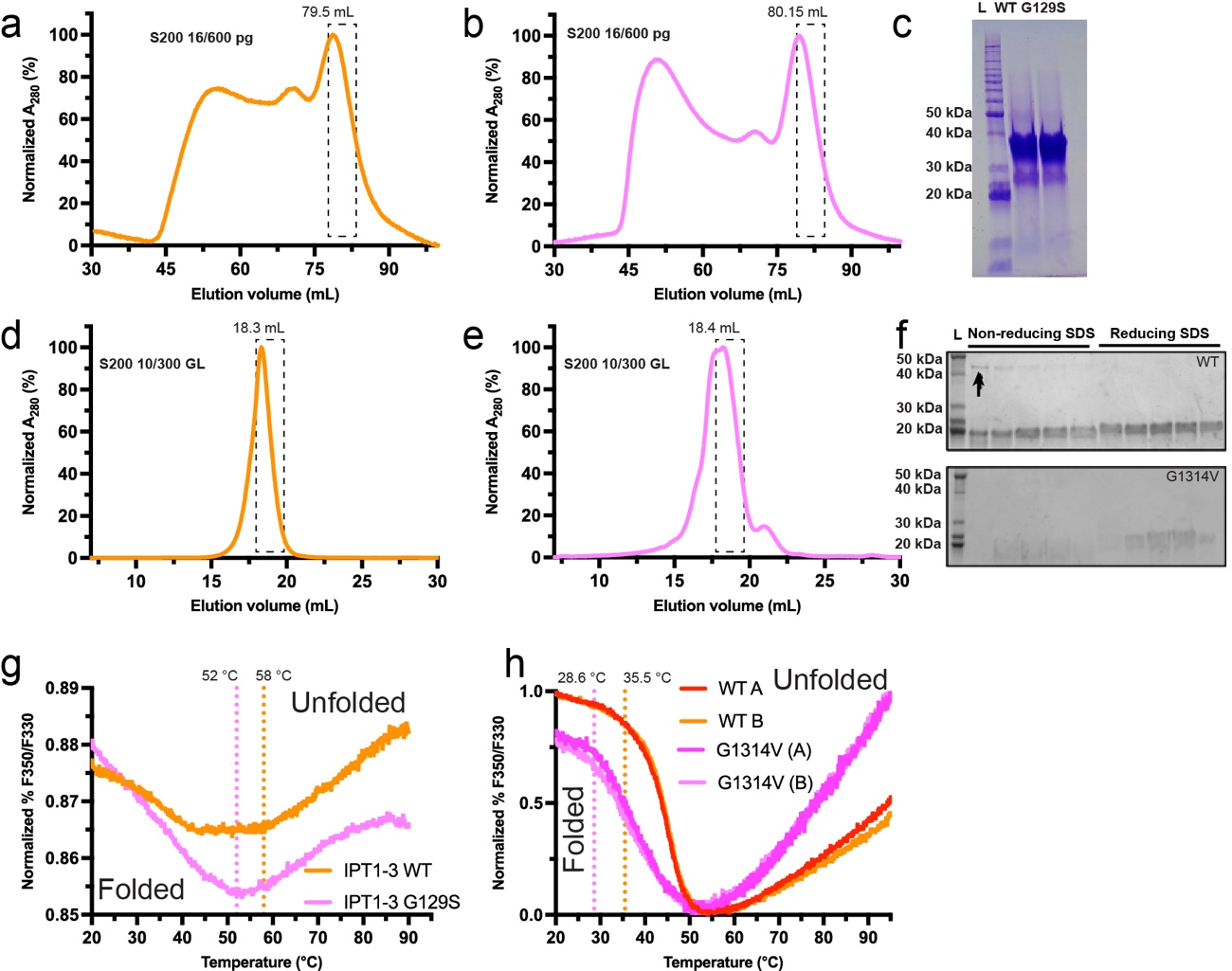

**Supplementary Fig. S5.** PKHD1L1 fragment protein purification and NanoDSF experiments.

PKHD1L1 protein fragments were purified using size exclusion chromatography. Absorbance (A_280_) was normalized and plotted as a function of elusion volume (mL). Next, purified fragment sample replicates were used for NanoDSF experiments. (a) SEC results for WT *Mm* PKHD1L1 IPT1-3 with elution peak at 79.5 mL. (b) SEC curve of *Mm* PKHD1L1 IPT1-3 p.(Gly129Ser) variant (elution peak at 80.5 mL). (c) Coomassie stained SDS-PAGE of eluted fractions from panels *a* and *b.* (d) SEC curve of WT *Mm* PKHD1L1 5-6 fragment. (e) SEC curve of *Mm* PKHD1L1 IPT5-6 fragment carrying the p.(Gly1314Val) variant. (f) Coomassie stained SDS-PAGE of eluted fractions from panels *d* and *e.* Both non-reducing and reducing SDS loading dyes (with and without *β*-mercaptoethanol) were used to determine the presence of artificial disulphide bond formation during refolding. A minor concentration of dimeric WT IPT5-6 was found, see black arrow in top panel. The dashed boxes in panel *a-e* indicate the SEC fractions used for NanoDSF measurements. (g) T_onset_ (melting temperature at which unfolding begins) NanoDSF measurement results presented as a function of normalized fluorescence ratio. WT *Mm* PKHD1L1 IPT1-3 (orange) and *Mm* PKHD1L1 IPT1-3 p.(Gly129Ser) variant (pink) are shown. Fluorescence intensity spectra shapes suggest a redshift unfolding pattern (https://2bind.com/nanodsf/). These results are related to Figure 4a. (h) T_onset_ measurement results for WT *Mm* PKHD1L1 IPT5-6 (red and orange) and p.(Gly1314Val) variant (purple and pink). Results suggest a blueshift unfolding pattern (https://2bind.com/nanodsf/). These results are related to Figure 4b. Dashed lines represent the temperatures at which unfolding begins.

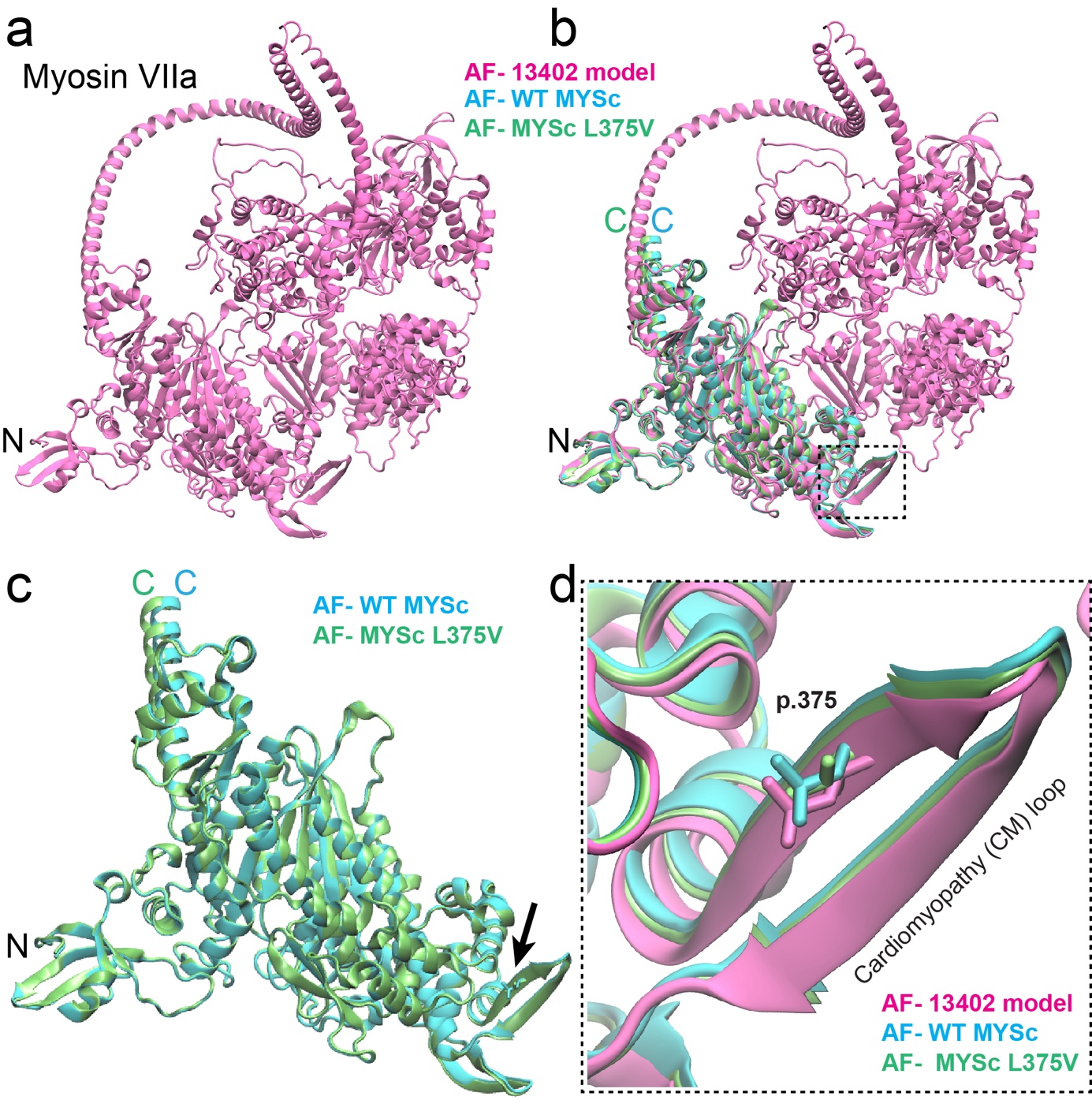

**Supplementary Fig. S6. AlphaFold2 modelling of human MYO7A** **protein.** (a) Alphafold2 model of the complete *Hs* MYO7A protein (mauve) extracted from the AlphaFold Protein Structure Database (entry code AF-Q13402-F1). (b) WT *Hs* MYO7A MYSc N-terminal domain (cyan), and the *Hs* MYO7A MYSc p.Leu375Val variant (lime) protein fragments were modeled using Alphafold2, and aligned with the AF-Q13402-F1 model (mauve). The dashed box outlines the region containing the cardiomyopathy (CM) loop carrying the MYO7A MYSc p.Leu375Val variant reported in Family 3. (c) Protein structural alignment of both the WT *Hs* MYO7A MYSc N-terminal domain (cyan) and *Hs* MYO7A MYSc p.Leu375Val variant (lime) as in panel *b,* comprising the residues p.1Met—Arg756 (NCBI code NP_000251.3). A clear overlap of the core MYSc domain is evidenced suggesting that AlphaFold2 is not able to predict any structural changes due to the p.Leu375Val variant (black arrow). (d) Zoom view from panel *b* (dashed box)*.* The CM loop shows a clear structural superposition following protein structural alignment. The WT MYO7A AF-Q13402-F1 (mauve) and WT MYSc domain (cyan) shows the side chains of the p.Leu375 residue. The p.V375 variant displayed in lime does not disrupt the folding feature of the CM β-strand. Since the amino acid residue change from p.Leu375 to p.Val375 imply a similar hydrophobicity in the CM region, and AlphaFold2 is not able to identify any plausible structural change, it is difficult to assess whether the p.Leu375Val mutation will be detrimental at both the structural and protein functional levels. This suggest that it is unclear if the p.Leu375Val variant will affect the structural properties of the CM loop or whether it will be potentially responsible for the functional relevance of the MYSc domain in MYO7A. The figures were rendered with the VMD software (Humphrey et al. 1996).

**Supplementary Table S1.**

| **Table S1.** List of species and PKHD1L1 accession numbers used for multiple sequence alignment analysis | | |
| --- | --- | --- |
| **Species** | **Abbreviation** | **NCBI Accession number** |
| *Homo sapiens****^&^*** | *Hs* | NP_803875.2 |
| *Otolemur garnettii* | *Og* | XP_023373972.1 |
| *Bos taurus* | *Bt* | XP_024857633.1 |
| *Sus scrofa* | *Ss* | XP_020944671.1 |
| *Canis lupus familiaris* | *Clf* | XP_038540795.1 |
| *Mus musculus* | *Mm* | NP_619615.2 |
| *Macaca mulatta* | *Mm2* | XP_015001362.2 |
| *Mustela lutreola* | *Ml* | XP_059021749.1 |
| *Pan troglodytes* | *Pt* | XP_016815270.3 |
| *Danio rerio* | *Dr* | NP_001305057.1 |

***^&^*** Variants were annotated using the *PHKD1L1* NM_177531.6 and ENST00000378402.9 accessions, corresponding to ENSP00000367655 and NP_803875.2. Signal peptides vary across the 10 PKHD1L1 orthologs (20 amino acids are predicted for *Hs* PKHD1L1 according to SMART).

**Supplementary Table S2.**

**Table S2.** Other variants excluded in the probands of Families 2, 3 and 4

|  |  |  |  |  | Max. MAF gnomAD (v3.1.2) | | Max. MAF gnomAD (v2.1.1) | | TOPMed v8 | All-of-Us |  |  |  |  |  |  | ACMG Criteria/Classification |  |
| --- | --- | --- | --- | --- | --- | --- | --- | --- | --- | --- | --- | --- | --- | --- | --- | --- | --- | --- |
| Gene | Transcript | cDNA (c.) | Protein Change (p.) | Zygosity | AF | AF Population | AF | AF Population |  |  | SIFT 2 | PolyPhen-2 | FATHMM | Mutation Taster | REVEL | CADD |  | Segregation |
| *LOXHD1* | NM_144612.6 | c.3124G>A | p.(Val1042Ile) | Het | 3.9e-4 | Latino/ Admixed American | 1.1e-3 | Latino/ Admixed American | 7.6e-5 | 1.4e-4 | 0.00^1^ | 0.99^1^ | 1.67^3^ | 0.105^3^ | 0.213^3^ | 22.4^1^ | PM2_P/VUS | Paternally inherited |
| *LOXHD1* | NM_144612.6 | c.5024G>A | p.(Arg1675His) | Het | 1.1e-3 | African/ African American | 6.3e-4 | African/ African American | 4.0e-4 | 2.7e-4 | 0.17^3^ | 0.998^1^ | -1.25^2^ | 0.808^3^ | 0.245^3^ | 22.8^1^ | PM2_P/VUS | Paternally inherited |
| *PCDH15* | NM_001354429.2 | c.2906A>T | p.(Asp969Val) | Het | 0 | -- | 0 | -- | -- | -- | 0.075^3^ | 0.037^3^ | 1.98^3^ | 0.708^3^ | 0.114^3^ | 23.3^1^ | PM2_P, BP4_P/VUS | Maternally inherited |
| *PCDH15* | NM_001354429.2 | c.1007G>A | p.(Arg336Gln) | Het |  | Other | 4.4e-4 | East Asian | 1.1e-5 | 1.8e-5 | 0.43^3^ | 0.025^3^ | 1.15^3^ | 1^1^ | 0.02^3^ | 22.4^1^ | PM2_P, BP4_P/VUS | Maternally inherited |
| *REST* | NM_005612.5 | c.2350C>T | p.(Pro784Ser) | Het | 0 | -- | 0 | -- | -- | -- | 0.24^3^ | 0.002^3^ | 3.17^3^ | 1^1^ | 0.016 ^3^ | 0.43 | PM2_P, BP4_P/VUS | Maternally inherited |
| *MYO7A* | NM_000260.4 | c.1123C>G | p.Leu375Val | Hom | 4.1e-4 | South Asian | 2.4e-4 | South Asian | -- | 3.4e-5 | 0.025^1^ | 0.08^3^ | -2.18^1^ | 1^1^ | 0.31^3^ | 21.5^1^ | PM2_P/VUS | Not Applicable |
| *POLD1* | NM_001256849.1 | c.1837G>A | p.Ala613Thr | Het | 0 | -- | 0 | -- | 1.1e-5 | 2.0e-6 | 0^1^ | 0.999^1^ | 2.00^3^ | 1^1^ | 0.577^2^ | 6.96^3^ | PM2_P/VUS | Maternally inherited |

Pathogenicity is represented as ^1^deleterious, ^2^neutral, or ^3^benign prediction, whereas “-” represents variant not scored.

Abbreviations: VUS, variant of uncertain significance
